# Type I interferon drives dysfunction of a distinct CD8+ HLA-DRB1+ T cell subset in systemic lupus erythematosus

**DOI:** 10.64898/2026.01.02.26343348

**Authors:** Huizhong Long, Elio Carmona, Mikhail G Dozmorov, Amr H Sawalha

**Affiliations:** Department of Orthopedics, Xiangya Hospital, Central South University, Changsha, Hunan, China; Division of Rheumatology, Department of Pediatrics, Children’s Hospital of Pittsburgh, University of Pittsburgh, Pittsburgh, Pennsylvania, USA; Department of Biostatistics, Virginia Commonwealth University, Richmond, Virginia, USA; Department of Pathology, Virginia Commonwealth University, Richmond, Virginia, USA; Division of Rheumatology and Clinical Immunology, Department of Medicine, University of Pittsburgh, Pittsburgh, Pennsylvania, USA; Lupus Center of Excellence, University of Pittsburgh School of Medicine, Pittsburgh, Pennsylvania, USA; Department of Immunology, University of Pittsburgh, Pittsburgh, Pennsylvania, USA

**Keywords:** T cells, lupus, CD8 cells, CD8+ HLA-DRB1+ T cells, cytotoxicity, interferon, exhaustion, senescence

## Abstract

**Objective:** Systemic lupus erythematosus (SLE) is characterized by persistent type I interferon (IFN) signaling and adaptive immune dysregulation. We previously identified hypomethylation of *HLA-DRB1* and *STAT1* in SLE CD8+ T cells, enabling aberrant IFN-driven HLA-DRB1 expression and expansion of a distinct CD8+ T cell subset. This study aimed to comprehensively characterize CD8+ HLA-DRB1+ T cells in lupus.

**Methods:** Peripheral blood CD8+ T cells from SLE patients and healthy controls were analyzed by flow cytometry to assess differentiation and effector functions. Single-cell RNA sequencing and TCR sequencing, with and without IFN-α stimulation, were used to assess transcriptional heterogeneity, exhaustion, senescence, and cytotoxicity.

**Results:** CD8+ HLA-DRB1+ T cells were enriched within effector memory and terminally differentiated CD8+ T cells and were significantly expanded within the effector memory compartment in SLE compared to healthy controls. These cells displayed paradoxical features of cytotoxic activation, proliferative potential, exhaustion, and senescence. Compared to healthy controls, lupus CD8+ HLA-DRB1+ T cells exhibited increased exhaustion, reduced cytotoxicity, and impaired viral defense pathways. IFN-α treatment enhanced IFN-γ responses in lupus CD8+ HLA-DRB1+ T cells and exacerbated exhaustion and senescence. Despite upregulation of cytotoxic gene expression, IFN-α reduced CD107a surface mobilization, indicating impaired degranulation. Analysis of lupus nephritis datasets revealed that most kidney-infiltrating CD8+ T cells express HLA-DRB1.

**Conclusion:** CD8+ HLA-DRB1+ T cells represent a cytotoxic yet dysfunctional effector memory population expanded in SLE. Type I IFN drives this paradoxical state by promoting exhaustion and impaired degranulation, highlighting a potential therapeutic axis in SLE.

## Introduction

Systemic lupus erythematosus is a chronic relapsing autoimmune disease characterized by the presence of autoantibodies and excessive type-I interferon production. Genetic, and environmental factors are involved in the pathogenesis of lupus ^1, 2^. Further, epigenetic mechanisms, and in particular DNA methylation changes, have been shown to play a role in the disease etiology ^3^.

While T cell DNA methylation studies in lupus have primarily focused on CD4+ T cells, epigenetic dysregulation in lupus CD8+ T cells has not previously been comprehensively explored. CD8+ T cells play a vital role in controlling viral infections, which may contribute to disease flares in lupus through enhanced type I interferon production. Epstein-Barr virus (EBV) infection, in particular, has been implicated in the pathogenesis of lupus, with evidence linking it to elevated type I interferon levels and, more recently, to underlying genetic susceptibility in lupus ^4–8^. Additionally, impaired cytotoxic function in a subset of CD8+ T cells has recently been associated with increased susceptibility to infections in lupus patients. Indeed, dysregulated CD8+ T cell responses have been observed across several autoimmune diseases ^9^. In lupus, a reduction in CD8+ effector memory T cells has been reported ^10, 11^, indicating that therapeutic approaches aimed at expanding this T cell subset may help mitigate virus-driven type I interferon production and subsequent disease flares ^11^. In addition, a subset of CD8+ T cells with impaired cytotoxic function has been associated with higher infection rates in patients with lupus. ^12, 13^. Recent data revealed the presence of activated CD8+ T cells in the kidneys of patients with lupus ^14^.

We previously reported marked hypomethylation at CpG sites in *HLA-DRB1* and *STAT1* in lupus CD8+ T cells compared to healthy controls ^15^. Given that interferon/STAT1 signaling promotes MHC class II expression, and that type I interferon levels are elevated in lupus, we hypothesized that hypomethylation at these loci may enable aberrant HLA-DRB1 expression on lupus CD8+ T cells in response to type I interferon. Indeed, our previous work revealed surface expression of HLA-DRB1 on a subset of CD8+ T cells that is expanded in lupus patients compared to healthy controls. Moreover, type I interferon induces HLA-DRB1 expression in lupus CD8+ T cells, but not in those from healthy individuals. These findings suggest that epigenetic priming enables interferon-driven HLA-DRB1 overexpression in lupus CD8+ T cells ^15^.

In this study, we sought to comprehensively characterize the phenotype, differentiation state, transcriptional profile, and functional properties of CD8+ HLA-DRB1+ T cells in lupus. Using a combination of flow cytometry and single-cell RNA sequencing, we defined the landscape of CD8+ T cell heterogeneity in lupus patients and healthy controls. Further, we investigated how this landscape is altered by type I interferon exposure, and explored the developmental trajectories and gene regulatory programs of key CD8+ T cell subsets. Our findings highlight a paradoxical population of CD8+ HLA-DRB1+ T cells that simultaneously exhibits features of cytotoxic activation, exhaustion, and senescence, and suggest that this population may be shaped by chronic type I interferon exposure in lupus.

## Methods

### Sample collection and cell separation

Blood samples were obtained from lupus patients and healthy controls after obtaining an informed written consent approved by the institutional review board of the University of Pittsburgh. Peripheral blood mononuclear cells (PBMCs) were isolated from whole blood using Ficoll-Paque density gradient centrifugation. PBMCs were immediately used for CD8+ T cell isolation and subsequent flow cytometry, or were cryopreserved in liquid nitrogen in a medium containing 10% dimethylsulfoxide and 90% fetal bovine serum for further experiment and scRNA-seq library construction and sequencing.

### CD8+ T cell isolation, stimulation, and treatment

For flow cytometry, CD8+ T cells were purified with negative selection from freshly isolated PBMCs using Miltenyi Biotec CD8+ T cell isolation kit following the manufacturer’s instructions. CD8+ T cells were then divided into two portions to study unstimulated and stimulated cells. For CD8+ T cells stimulation, cells were stimulated overnight in cell culture wells precoated with 10 µg/mL of anti-human CD3 (Clone: UCHT1) and 2.5 µg/mL of soluble anti-human CD28 (Clone: 28.2) (BD Biosciences) in T cell media (RPMI media with 10% fetal bovine serum, 2 mM L-glutamine, 100 U/mL penicillin, and 100 mg/mL streptomycin). Culture media were then replaced with media without anti-human CD3 or CD28, and cells were cultured for an additional 4 days. Unstimulated CD8+ T cells were cultured in the same way but without anti-human CD3/CD28.

For scRNA-seq, CD8+ T cells were purified with negative selection from thawed PBMCs using Miltenyi Biotec CD8+ T cell isolation kit following the manufacturer’s instructions. CD8+T cells were stimulated with anti-human CD3/CD28 as described above, with or without IFN-α (1000 IU/mL).

### Flow cytometry

For the last 6 hours of incubation, CD8+ T cells were incubated with surface staining anti-CD107a-APC mAb (H4A3, BioLegend). CD8+ T cells were also incubated with cytokine secretion blocker Brefeldin A solution (Golgi plug, 5 lg/ml; BioLegend) and Monensin (Golgi stop, 1 lL/ml; eBioseicence) for the last 4 hours. After incubation, CD8+ T cells were stained with Live/Dead Fix Near IR dead cell stain kit (Invitrogen, L10119) in the dark for 20 min at room temperature. The surface antibody cocktail (CD3: QDOT605, UCHT1, Thermo Fisher Scientific; CD8: BUV395, RPA-T8, BD Biosciences; CD45RA: BV510, HI100, BioLegend; CCR7: G043H7, BV650, BioLegend; HLA-DRB1: NFLD.D2, PE, BioLegend; CD319: FITC, 162.1, BioLegend) was added to pre-stained cells and incubated for 20 min at 4℃. CD8+ T cells were then fixed, and permeabilized using Fixation Buffer and Intracellular Staining Permeabilization Wash Buffer (both from BioLegend) before intracellular staining. The intracellular antibody cocktail (GZMA: R718, CB9, BD Biosciences; GZMB: PE-Dazzle 594, QA16A02, BioLegend; Perforin: BV421, B-D48, BioLegend; IFN-γ: BV750, 4S.B3, BioLegend) was added to CD8+ T cell suspensions and incubated for 20 min at 4℃ in the dark. Flow cytometry data were acquired using the LSRFortessa cell analyzer and analyzed using FlowJo software version 10.8 (both from BD Biosciences). The gating strategy for identifying CD8+ HLA-DRB1+ T cells is shown in **Supplementary Figure 1**.

### Single-cell RNA sequencing and single-cell TCR sequencing

Isolated CD8+ T cell suspensions from 6 SLE patients and age-, sex-, and race-matched healthy controls were incubated with a unique TotalSeq-C anti-human Hashtag antibody conjugated to a Feature Barcode oligonucleotide (BioLegend) for multiplexing. After washing, cell suspensions were pooled and used to generate single-cell gel beads in emulsion. A single-cell 5’ gene expression sequencing library, paired single-cell T-cell receptor (TCR), and cell surface protein Feature Barcode library were prepared following 10X Genomics instructions in the Single Cell Core at the University of Pittsburgh. Libraries were sequenced using Novaseq 6000 to achieve 25,000 reads per cell for the gene expression library, 5,000 reads for the TCR library, and 5,000 reads for the Feature Barcode library.

### Single-cell data processing

Sample demultiplexing, barcode processing, and single-cell counting were performed using Cell Ranger (Version 7.1.0, 10X Genomics). Cell Ranger cell count was used to align samples to the GRCh38 reference genome to generate a raw gene expression matrix for each sample. TCR-seq reads were aligned to the human (GRCh38) V(D)J reference to assemble TCR sequences. Downstream single-cell analysis was carried out with R (version 4.2.0) using the Seurat package (version 5.2.0) ^16^. TCR sequencing data were analyzed using the scRepertoire package ^17^. Single cells were excluded if they expressed <600 unique genes, >6,000 unique genes, or expressed more than 10% mitochondrial DNA content. Genes appearing in less than three cells were also excluded. Filtered data were normalized using a scaling factor of 10,000. The 2,000 highly variable genes were selected using the FindVariableFeatures function. The expression of each gene was then scaled by the ScaleData function. Data from different datasets were then merged into one Seurat subject. Principal component analysis (PCA) was performed using the RunPCA function with default parameters. Then the Seurat object was integrated using the R package Harmony ^18^ with datasets as the grouping variable to correct for batch effects.

### Clustering and annotation

A shared nearest-neighbor graph using the first harmony-adjusted 25 PCs was calculated, followed by clustering using the FindClusters function with resolution set to 1.0. Graphical projections of these cluster analysis results were constructed via the Uniform Manifold Approximation and Projection (UMAP) methodology. The FindAllMarkers function implemented in Seurat was used to identify the differentially expressed genes within each subset. Clusters were then manually annotated based on cluster-specific differentially expressed genes. Selected clusters were merged to represent major cell types. Clusters with low quality scores were excluded from further analyses.

### Differential expression and functional enrichment analyses

Differentially expressed genes (DEG) between two groups or different cells were determined using the original expression data in RNA assay via the FindMarkers function in the Seurat package based on two-sided Wilcoxon rank-sum test. Genes that met the criteria of adjusted P-value <0.05 (using Benjamini-Hochberg correction) were considered significant DEGs with parameters min.pct = 0.1 and logfc.threshold = 0.25. Gene Ontology (GO) and Kyoto Encyclopedia of Genes and Genomes (KEGG) enrichment analyses of DEGs were performed using the R package clusterProfiler ^19^.

### CD8+ T cell signature scores

CD8+ T cell signature scores were calculated per cell using the AddModuleScore function from Seurat. Related genes were used for calculating exhaustion, cytotoxicity, and inflammatory scores according to published T cell studies ^20–22^.

## Results

### Differentiation status of CD8+ HLA-DRB1+ T cells in SLE

We previously identified a unique CD8+ T cell subset co-expressing HLA-DRB1 in patients with SLE^15^. To determine the differentiation status of this subset (naïve, memory, or effector), we performed multi-parameter flow cytometry. Our analysis revealed that, ex vivo, CD8+ HLA-DRB1+ T cells span the effector memory (EM), terminally differentiated effector memory (TEMRA), and naïve CD8+ T cell compartments. Upon CD3/CD28 stimulation, the majority of naïve CD8+ HLA-DRB1+ T cells differentiate into EM cells in both SLE patients and healthy controls. In stimulated and unstimulated cells, there was a trend for more TEMRA cells and fewer EM cells in SLE patients compared to healthy controls though without statistical significance (**Figure 1**).

**Figure 1:**
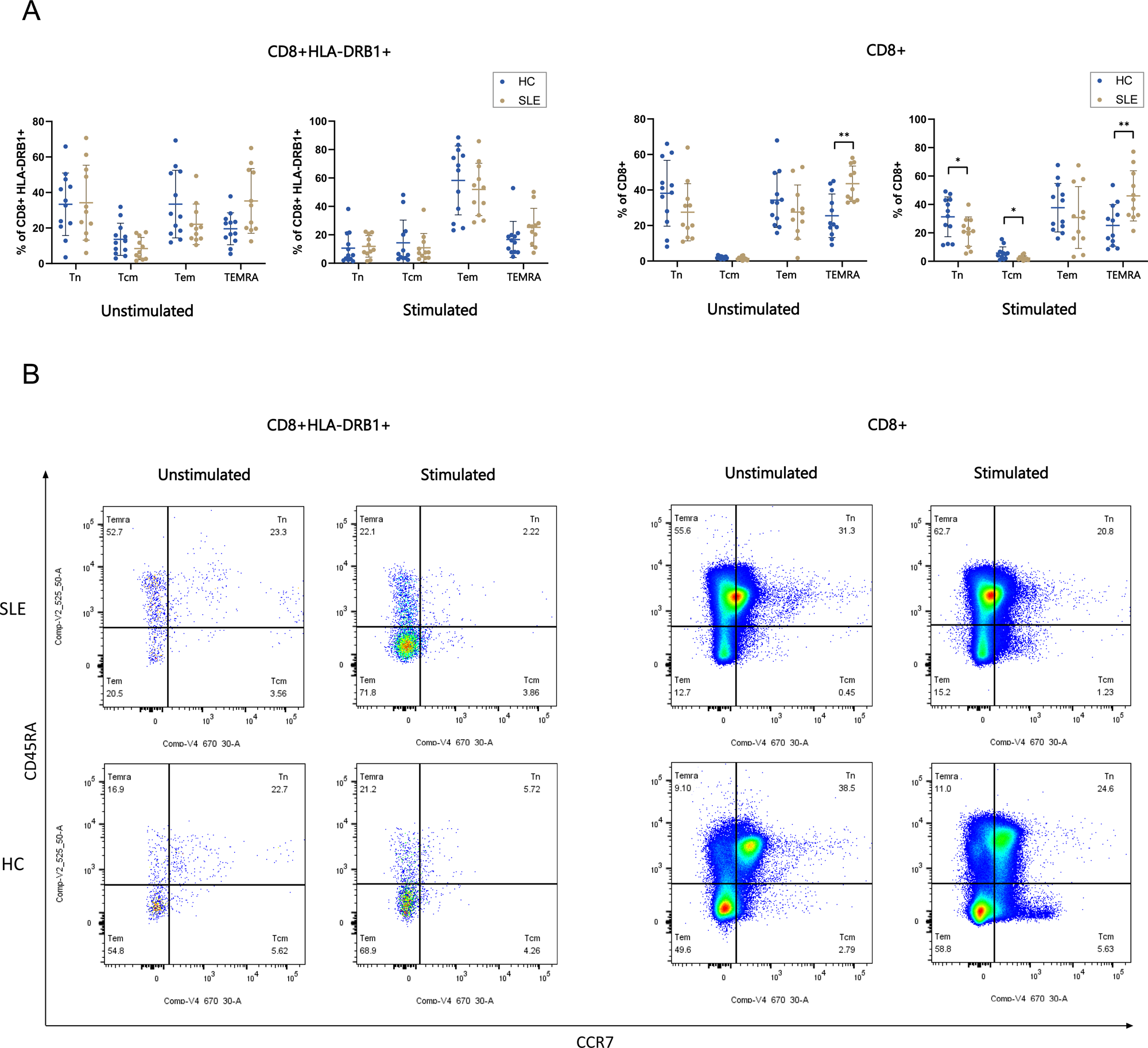
Flow cytometry analysis of CD8+ HLA-DRB1+ T cells and total CD8+ T cells in patients with SLE compared to healthy controls. **(A)** The percentage of naïve, central memory (CM), effector memory (EM) and terminally differentiated effector memory (TEMRA) cells were evaluated in CD8+ HLA-DRB1+ T cells (left panel) and all CD8+ T cells (right panel) from patients with SLE and healthy control (HC) with and without CD3/CD28 stimulation. *P < 0.05, **P < 0.01, 2-tailed Mann-Whitney Test. A representative dot plot is shown in panel **(B)**.

### Global CD8+ T cell subset skewing in SLE

To investigate broader CD8+ T cell dysregulation in SLE, we compared subset stratification across SLE patients and healthy controls. Our analysis revealed that compared to healthy controls, stimulated CD8+ T cells from SLE patients consist of significantly higher proportion of TEMRA cells and lower proportion of naïve and central memory (CM) CD8+ T cells. SLE patients had higher proportion of TEMRA CD8+ T cells ex vivo, prior to stimulation (**Figure 1**).

### Single cell transcriptomic and TCR profiling of CD8+ HLA-DRB1+ T cells

To further characterize the CD8+ HLA-DRB1+ T cell subset, we performed scRNA-seq and scTCR-seq in stimulated CD8+T cells from SLE patients and age-, sex-, and race-matched healthy controls. After quality control and unsupervised clustering, 6 cell types were identified, including naïve CD8+ T cells, CM CD8+ T cells, EM CD8+ T cells, TEMRA CD8+ T cells, proliferative CD8+ T cells, and Mucosal-Associated Invariant T cells (MAIT) (**Figure 2A, Supplementary Figure 2**).

**Figure 2.**
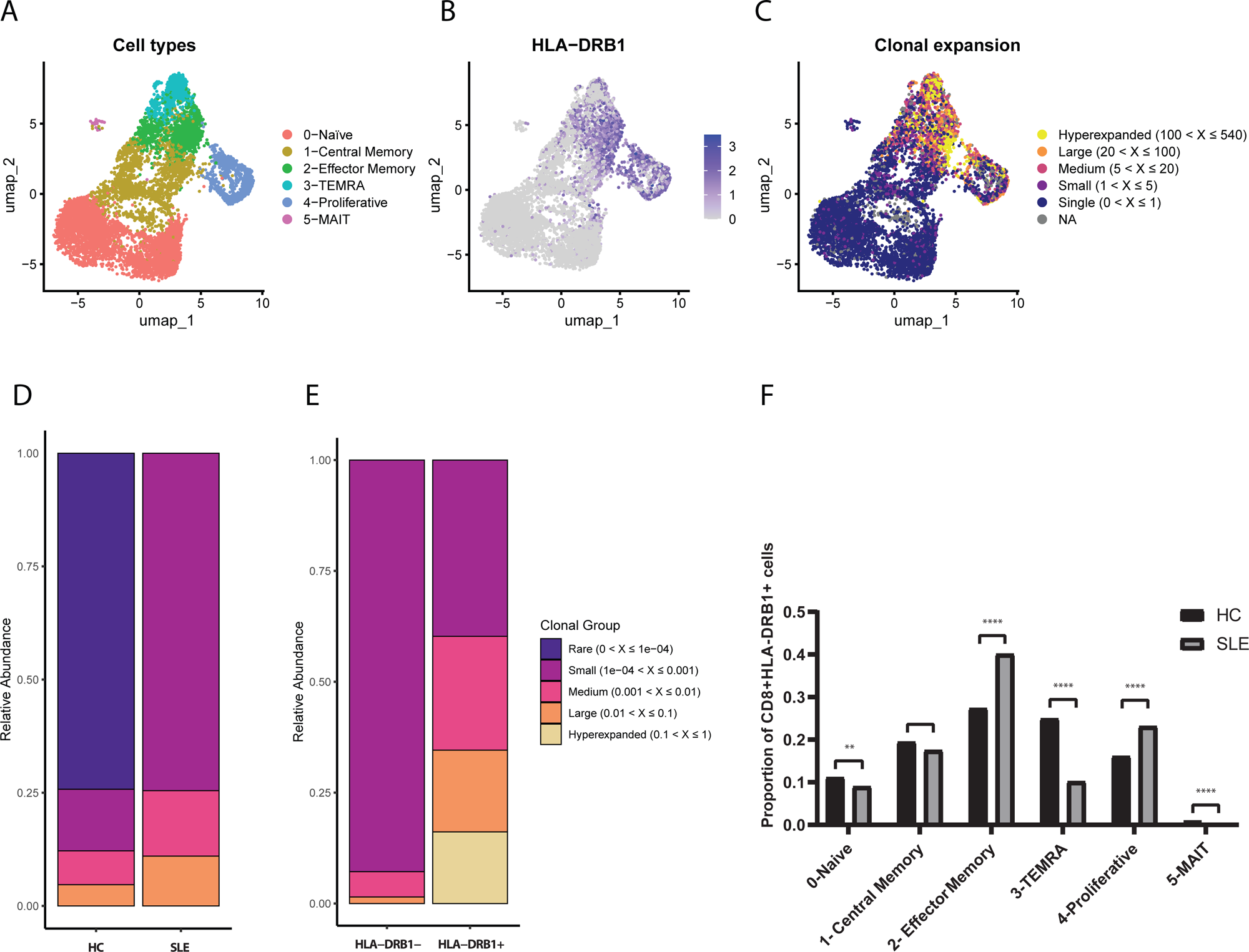
Single-cell RNA-seq and single-cell TCR-seq in CD3/CD28 stimulated CD8+ T cells from patients with SLE and healthy controls. **(A)** UMAP plots depicting the cell cluster subpopulations in SLE CD8+ T cell after CD3/CD28 stimulation. **(B)** Feature plot depicting the expression of HLA-DRB1 in SLE CD8+T cells based on UMAP. **(C)** UMAP showing the expansion of clonotypes in SLE CD8+ T cells. TCR clonotypes were classified according to their extent of clonal expansion. **(D)** Stacked bar plot showing the relative proportions of different cell expansion categories in CD8+ T cells from SLE patients and healthy controls. **(E)** Stacked bar plot showing the relative proportions of different cell expansion categories across in SLE HLA-DRB1+ and HLA-DRB1- CD8+ T cells. **(F)** Differences in the proportions of HLA-DRB1+ CD8+ T cell subpopulations between SLE and healthy controls (HC).). **P < 0.01, ****P < 0.0001, Chi-squared test.

Consistent with the flow cytometry data, following stimulation, CD8+ HLA-DRB1+ T cells were mainly effector-related cells including EM, proliferative T cells, and TEMRA CD8+ T cells, while CD8+ HLA-DRB1− T cells were primarily naïve and CM T cells (**Figure 2B**). Analysis using TCR repertoire revealed evidence for clonal expansion in lupus CD8+ T cells compared to healthy controls, which is particularly more evident in CD8+ HLA-DRB1+ T cells (**Figure 2C-E**). Compared to healthy controls, CD8+ HLA-DRB1+ T cells in lupus were most significantly expanded within EM CD8+ T cells, proliferative T cells, but reduced in TEMRA cells (**Figure 2F**). Indeed, over 40% of EM CD8+ T cells in SLE patients are HLA-DRB1+ (**Figure 2F**).

### Cytotoxic, exhaustion, and senescence signatures of CD8+ HLA-DRB1+ T cells

Compared to CD8+ HLA-DRB1- T cells, The CD8+ HLA-DRB1+ T cells in SLE patients demonstrated a significantly elevated cytotoxic signature, as evidenced by upregulated expression of genes encoding cytolytic mediators (e.g., GZMA, GZMB, GZMK, PRF1, SLAMF7, NKG7, GNLY), along with elevated inflammatory signature characterized by heightened expression of IFNG and the proliferation marker MKI67. Notably, this population exhibited higher exhaustion scores (with higher expression of LAG3, TIGIT) and senescence (upregulated CDKN2A [encoding p16, a senescence-associated cyclin-dependent kinase inhibitor], KLRG1, and loss of costimulatory molecules CD27/CD28). These data suggest a paradoxical coexistence of cytotoxic activation, proliferative potential, and terminal differentiation signals characterizing CD8+ HLA-DRB1+ T cells in SLE (**Figure 3, Supplementary Figure 3, Supplementary Table 1**). Flow cytometry results confirmed upregulation of cytotoxicity-related molecules and IFN-γ production in CD8+ HLA-DRB1+ T cells at the protein levels (**Supplementary Figure 4**). KEGG enrichment analyses were conducted to identify pathways enriched by genes upregulated in CD8+ HLA-DRB1+ T cells. The analyses revealed significant enrichment in pathways such as cellular senescence, apoptosis, and T cell receptor signaling pathway, among others (**Supplementary Figure 5**). Analysis of the characteristics of CD8+ HLA-DRB1+ T cells from healthy controls revealed similar results (**Supplementary Figure 6-9, Supplementary Table 2**).

**Figure 3:**
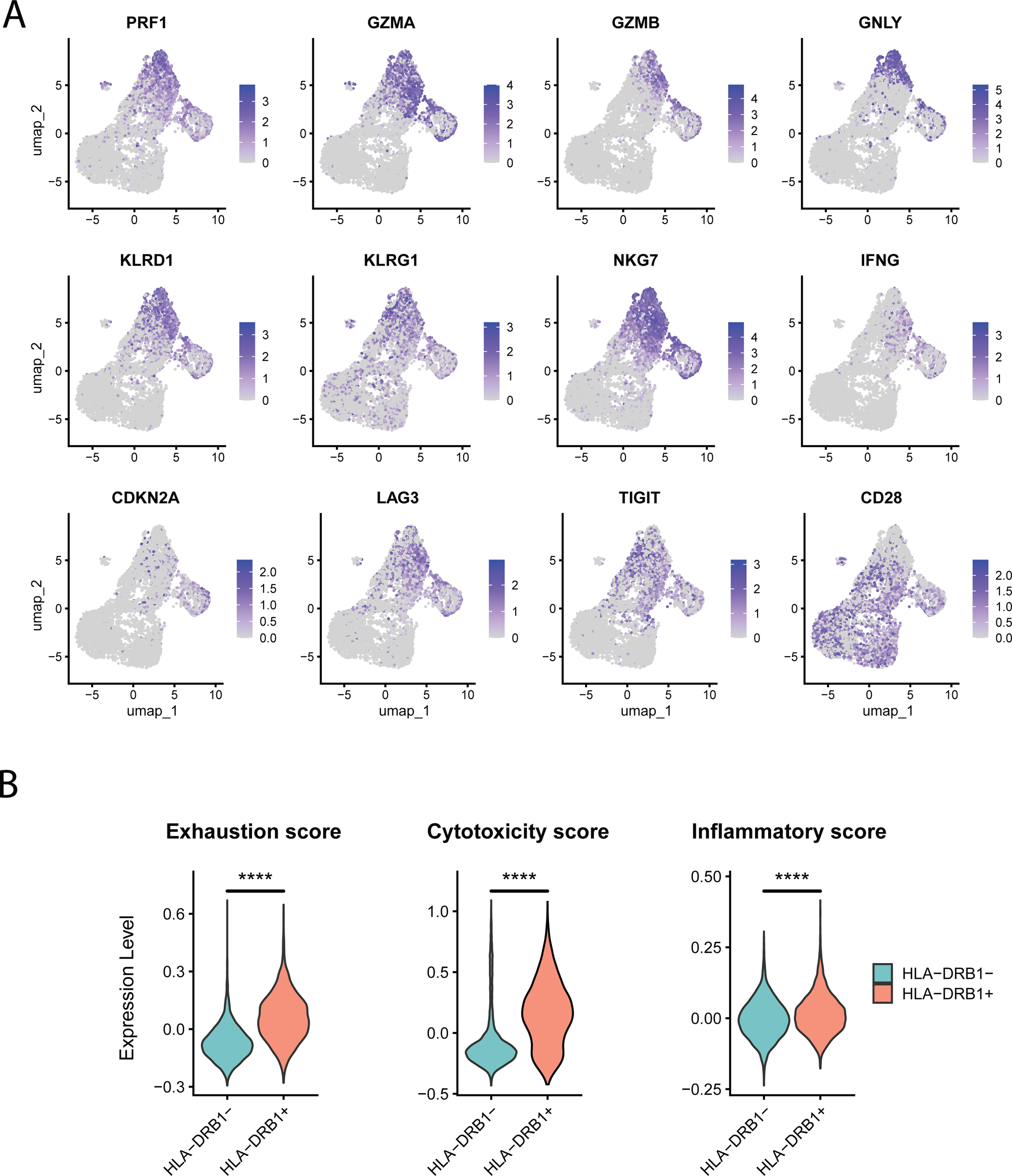
CD8+ HLA-DRB1+ T cells exhibit high cytotoxicity alongside features of exhaustion and senescence. **(A)** Feature plot depicting the expression of cytotoxicity, exhaustion and senescence markers in SLE CD8+T cells based on UMAP. **(B)** Violin plots comparing the exhaustion, cytotoxicity, and inflammatory scores in SLE CD8+ HLA-DRB1+ vs CD8+HLA-DRB- T cells. ****P < 0.0001, 2-tailed t test.

### Transcriptional differences between SLE and healthy control CD8+ HLA-DRB1+ T cells

Next, we compared CD8+ HLA-DRB1+ T cells between SLE patients and healthy controls. In SLE patients, this subset exhibited significantly reduced cytotoxicity score at the transcriptional level compared to healthy controls, along with higher exhaustion scores (**Figure 4A**). DEG analysis revealed upregulated expression levels of various genes associated with exhaustion and immunosuppression including ENTPD1, TNFAIP3, LAG3, EZH2, JUN, MAP3K5, MAPK13, LMNB1, and XIST, along with higher IFNG expression. Further, downregulation of S1PR5, and CX3CR1 in SLE HLA-DRB1+ CD8+ T cells were observed, indicating impaired cytotoxicity and dysregulated cell trafficking (**Supplementary Table 3**). Moreover, the proliferation marker MKI67 was also significantly overexpressed in HLA-DRB1+ CD8+ T cells from SLE patients compared to healthy controls (**Figure 4B**). Gene Ontology analysis revealed that defense response to viral infections was enriched among downregulated genes (**Figure 4C**), while processes such as chromosome segregation and organelle fission were enriched in upregulated genes (**Figure 4D**).

**Figure 4:**
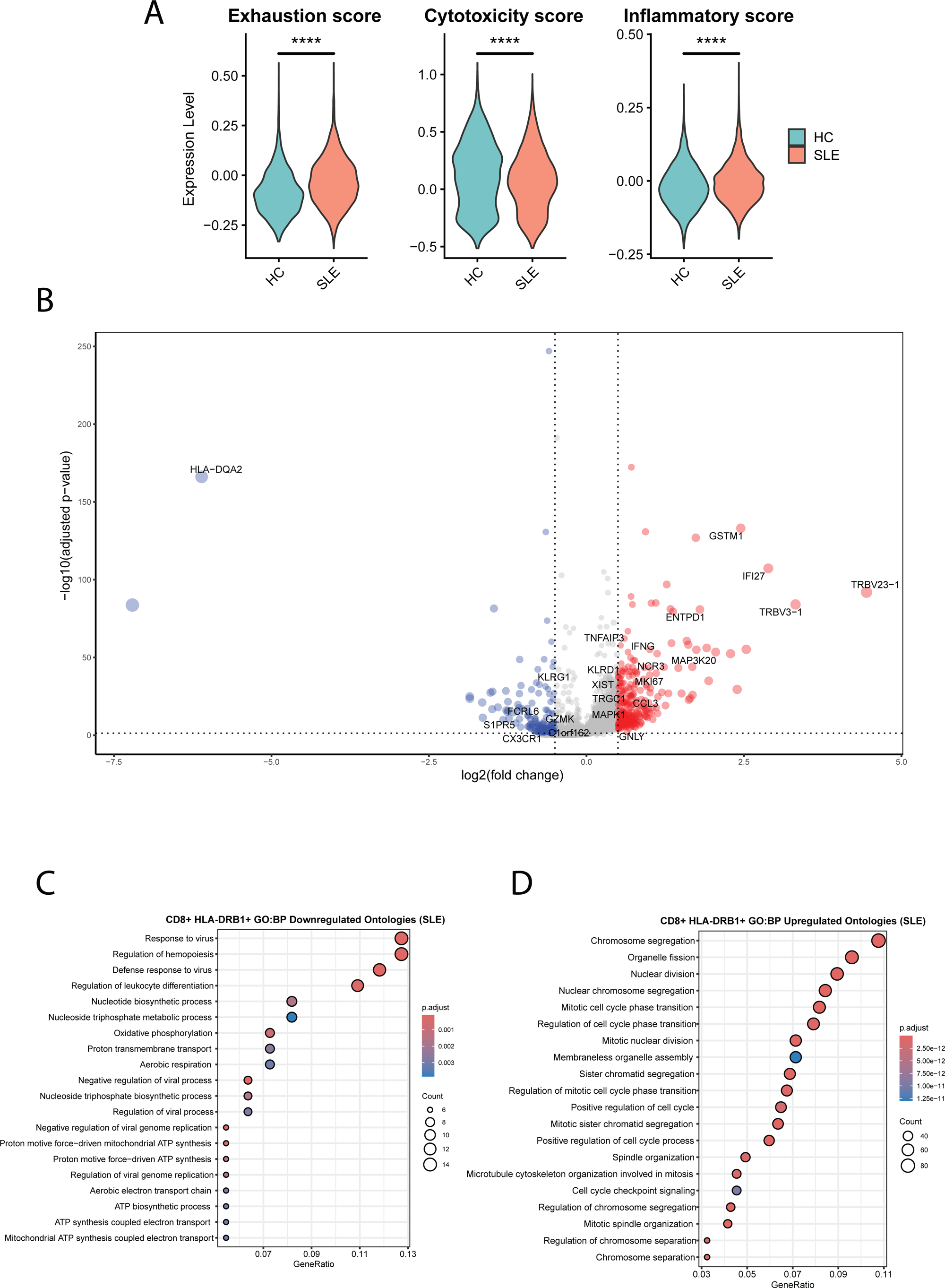
CD8+HLA-DRB1+ T cell from patients with SLE exhibited an exhausted and immunosuppressive phenotype. **(A)** Violin plots comparing the exhaustion, cytotoxicity, and inflammatory scores across CD8+ HLA-DRB1+ T cell from SLE patients and healthy controls (HC). ****P < 0.0001, 2-tailed t test. **(B)** A volcano plot illustrating the DEGs in CD8+ HLA-DRB1+ T cell from SLE patients and HC. Genes upregulated in SLE patients are shown in red and downregulated genes are shown in blue. **(C)** Representative Gene Ontology (Biological Process) terms enriched in genes downregulated in CD8+ HLA-DRB1+ T cell from SLE patients compared with HC. **(D)** Representative Gene Ontology (Biological Process) terms enriched in genes upregulated in CD8+HLA-DRB1+ T cell from SLE patients compared with HC.

### Effects of IFN-α on CD8+ HLA-DRB1+ T cells

We then determined the effect of IFN-α treatment on CD8+ HLA-DRB1+ T cells. After treatment with IFN-α, the expression of cytotoxicity-related genes in SLE CD8+ HLA-DRB1+ T cells was significantly enhanced, as indicated by increased cytotoxicity scores and elevated expression of cytotoxicity-related genes such as GZMB, PRF1, GNLY, and NKG7. Further, IFNG expression and the inflammatory score were significantly increased in CD8+ HLA-DRB1+ T cells from SLE after treatment with IFN-α. Concurrently, T cell exhaustion of CD8+ HLA-DRB1+ T cells was further exacerbated (with upregulation of LAG3), and the cells exhibited an increased propensity toward a senescent phenotype, as evidenced by upregulated senescence-associated markers (CDKN1A, CDKN2A) and downregulated CD27, CD28, and CD161. Increased expression of immunoregulatory genes, including TNFAIP3 and CD38 were also observed (**Figure 5A, Supplementary Figure 10, Supplementary Table 4**). Further, GO enrichment analysis showed that response to virus, regulation of response to biotic stimulus, regulation of innate immune response, and interferon-mediated signaling pathway were significantly upregulated (**Figure 5B**). In the KEGG pathway enrichment analysis, pathways such as NOD-like receptor signaling, apoptosis, necroptosis, RIG-I-like receptor signaling, graft-versus-host disease, and allograft rejection exhibited significant enrichment (**Figure 5C**). In healthy controls, IFN-α treatment exerted comparable effects on CD8+ HLA-DRB1+ T cells, though it failed to induce significant upregulation of CDKN1A, CDKN2A, IFNG, and GNLY expression (**Supplementary Figure 11, Supplementary Table 5**).

**Figure 5:**
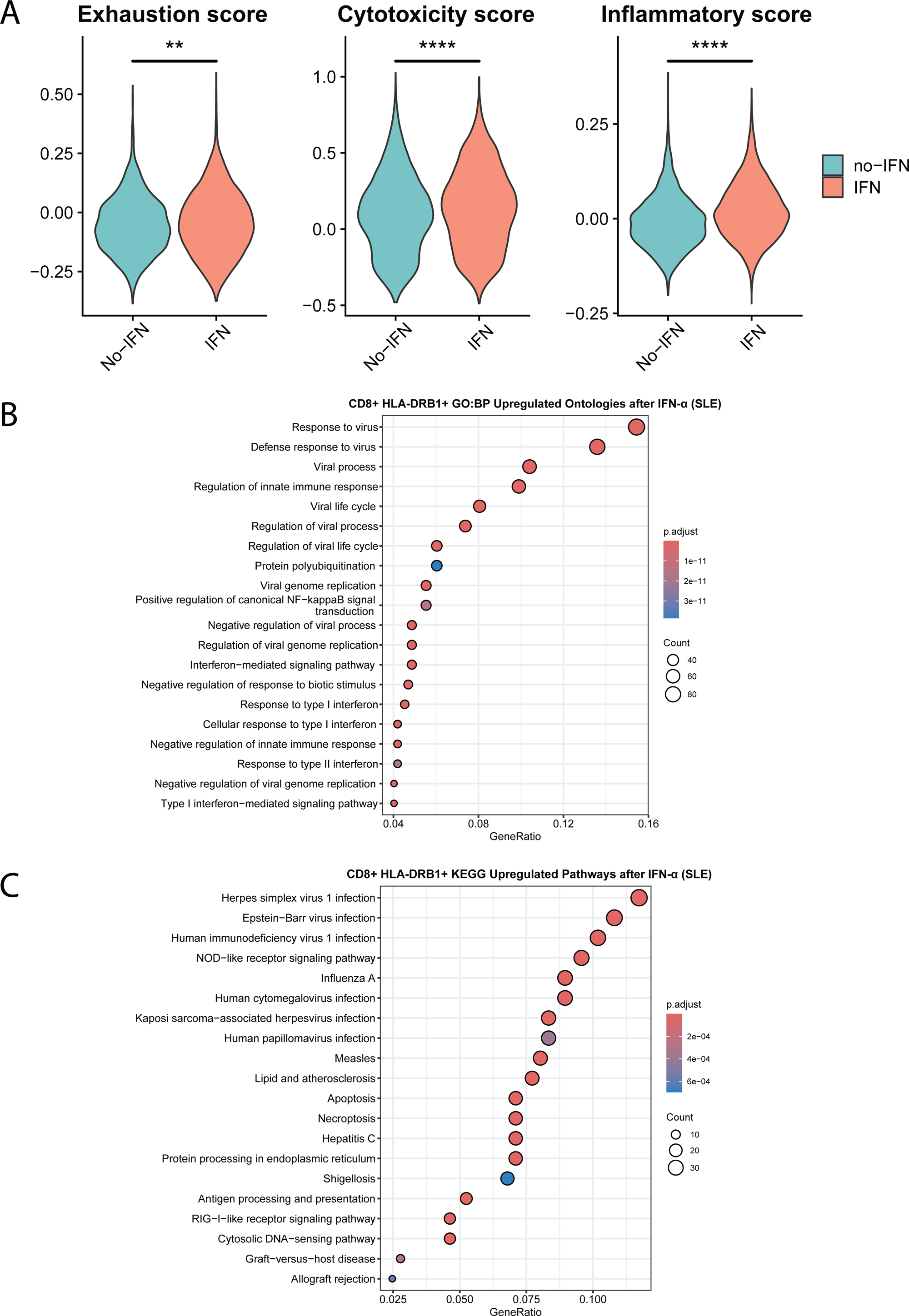
IFN-α treatment enhances the cytotoxicity of SLE CD8+ HLA-DRB1+ T cells but also promotes their transition toward exhausted and senescent phenotypes. **(A)** Violin plots comparing the exhaustion, cytotoxicity, and inflammatory scores in SLE CD8+ HLA-DRB1+ T cells with and without IFN-α treatment. **P < 0.01, ****P < 0.0001, 2-tailed t test. **(B)** Representative Gene Ontology (Biological Process) terms enriched in genes upregulated after IFN-α treatment in SLE CD8+ HLA-DRB1+ T cells. **(C)** Representative KEGG terms and pathways enriched in genes upregulated after IFN-α treatment in SLE CD8+ HLA-DRB1+ T cells.

### IFN-α induces dysfunctional cytotoxicity in CD8+ HLA-DRB1+ T cells

To examine if increased expression of cytotoxicity-related genes in SLE CD8+ HLA-DRB1+ T cells we observed with IFN-α is associated with increased cytotoxicity, we examined the expression of CD107a, as a degranulation marker, using flow cytometry. Flow cytometry confirmed that IFN-α treatment upregulated Perforin expression, a key cytotoxicity molecule, but was associated with reduced CD107a expression indicating impaired degranulation and therefore dysfunctional cytotoxicity (**Figure 6**). Indeed, CD8+ HLA-DRB1+ T cells from lupus patients had a similar trend of impaired degranulation as compared to healthy controls (**Supplementary Figure 12**).

**Figure 6:**
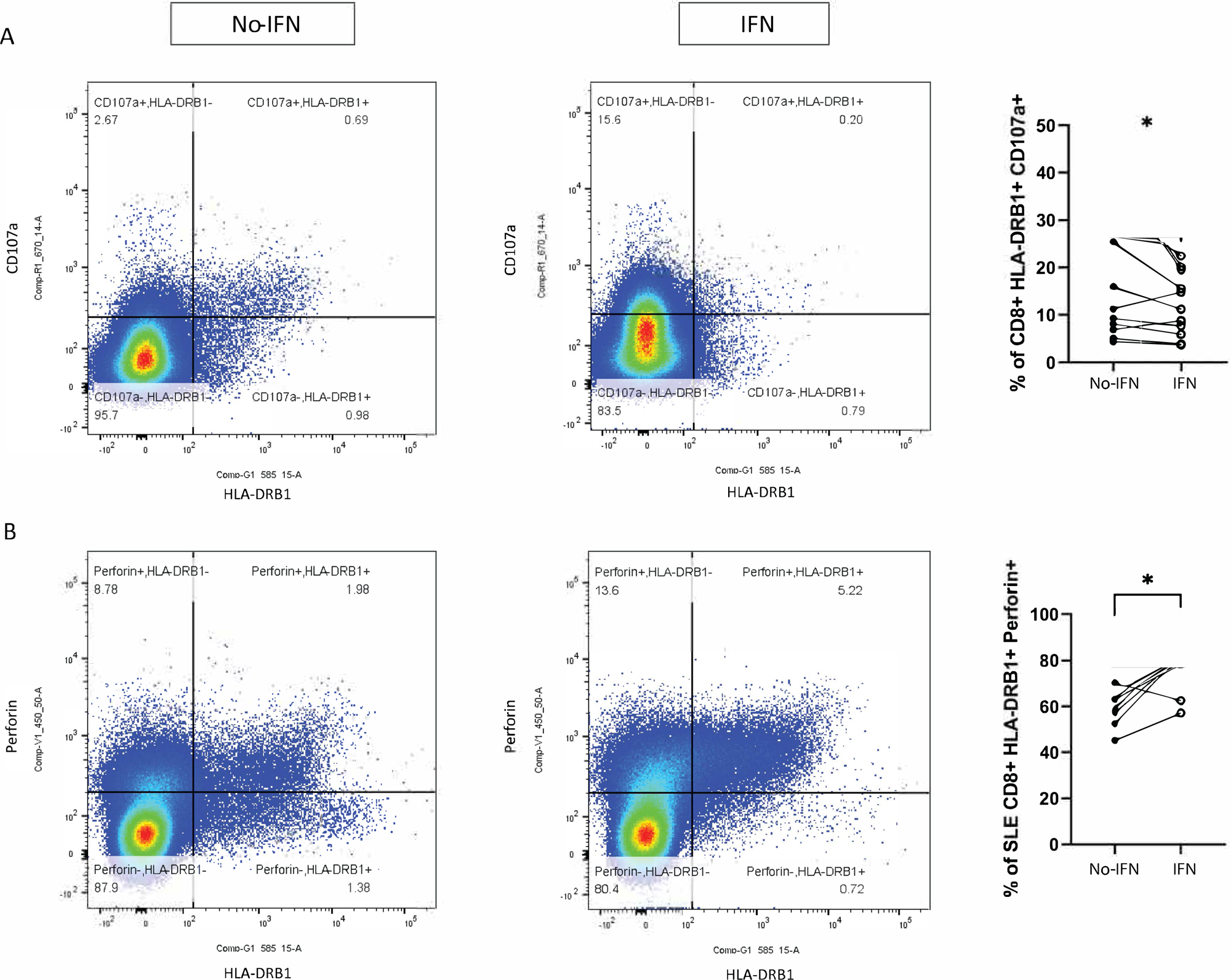
IFN-α treatment impaired degranulation, as reflected by reduced CD107a expression, and upregulated Perforin production in SLE CD8+ HLA-DRB1+ T cells. Representative flow cytometry plots and proportion of CD107a+ **(A)** and Perforin **(B)** in CD8+ HLA-DRB1+ T cells with and without IFN-α treatment are shown. *P < 0.05, Wilcoxon test.

### Effects of IFN-α on CD8+ HLA-DRB1- T cell subsets

We also examined the effect of IFN-α treatment on SLE CD8+ HLA-DRB1- T cells. After IFN-α treatment, a significant reduction was observed in the proportion of naïve CD8+ T cells, whereas the percentages of EM cells and TEMRA cells demonstrated marked increases (**Supplementary Figure 13A**). In EM T cells, IFN-α treatment induced upregulation of LAG3 and CD38, and downregulation of cytotoxicity-associated genes (e.g., GZMA, FGFBP2), with a concomitant reduction in cytotoxicity scores (**Supplementary Figure 13B, Supplementary Table 6**). IFN-α treatment also induced upregulation of LAG3, CD38, and downregulation of cytotoxicity scores in TEMRA cells, marked by significant lower expression of GZMA and FGFBP2 (**Supplementary Figure 13C, Supplementary Table 7**). The effects of IFN-α on CD8+ HLA-DRB1- T cell subsets from healthy controls were similar (**Supplementary Figure 14, Supplementary Tables 8 and 9**). Finally, we analyzed the effect of IFN-α treatment on total CD8+ T cells subset proportions in SLE patients and healthy controls. In both SLE and healthy controls, IFN treatment results in expansion of TEMRA cells and a reduction in naïve CD8+ T cells (**Supplementary Figures 15 and 16, Supplementary Tables 10 and 11**).

### Presence of CD8+ HLA-DRB1+ T cells in lupus nephritis kidneys

To further contextualize our findings in peripheral blood, we examined publicly available single-cell RNA-seq data from kidney tissue available from the Accelerating Medicines Partnership (AMP) lupus nephritis cohort. Based on these data, kidney biopsies from lupus patients demonstrated a notable presence of CD8+ T cells along with a clear interferon-response transcriptional signature ^14^. Three different CD8+ T cell subsets were reported in lupus kidneys ^14^. Consistent with our observations in peripheral blood, our analysis of the AMP dataset revealed expression of HLA-DRB1 in the majority of kidney-infiltrating CD8+ T cells (**Supplementary Figure 17**).

## Discussion

This study provides an in-depth characterization of a previously underexplored population of CD8+ T cells expressing HLA-DRB1 in systemic lupus erythematosus. Through a combination of flow cytometry, single-cell transcriptomics, and functional transcriptional measures, we demonstrate that CD8+ HLA-DRB1+ T cells in SLE are predominantly effector cells with distinct transcriptional and functional profiles, including enhanced cytotoxic potential, proliferative activity, and features of exhaustion and senescence. Importantly, we show that type I interferon (IFN-α) further accentuates the dysfunctional characteristics of this population, suggesting that persistent IFN signaling in SLE may exacerbate the paradoxical phenotype of these cells.

Importantly, flow cytometry data revealed that IFN-α impairs degranulation of CD8+ HLA-DRB1+ T cells in SLE, suggesting an overall interferon-induced dysfunctional cytotoxicity in these cells, despite an observed interferon-mediated upregulation of cytotoxic molecules. This interpretation is supported by our observation of reduced CD107a surface mobilization in CD8+ HLA-DRB1+ by IFN-α. Because CD107a (LAMP-1) is transiently expressed on the plasma membrane upon granule exocytosis, CD107a downregulation is widely used as a direct proxy for impaired cytotoxic cell degranulation and reduced delivery of perforin/granzymes to target cells^23^. Notably, reduced CD107a-defined degranulation in SLE CD8+ T cells has been previously reported and linked to disease activity^24^. This defect is enriched within a CD8+ CD38^high^ T cells, which also exhibited higher expression of HLA-DR, EZH2, and markers linked to cell activation and exhaustion^12^. Indeed, inhibiting EZH2 restored CD107a degranulation in CD8+ CD38^high^ T cells ^12^. Given our finding of increased EZH2 expression in lupus HLA-DRB1+ CD8 T cells, these data raise the possibility that IFN-driven activation programs in SLE may coexist with epigenetically constrained granule exocytosis, resulting in a cytotoxic “content-high but delivery-impaired” phenotype. Further, CD8+ HLA-DRB1+ T cells overexpress IFN-γ, with significantly higher levels in lupus patients compared to healthy controls, and this expression is further enhanced by type I interferon treatment.

IFN-γ production from CD8+ T cells and effector CD8+ T cell functions have been linked to glomerulonephritis and organ damage in lupus ^25, 26^. CD8+ T cells have been shown to be a predominant cell type infiltrating renal tissue in patients with lupus nephritis ^27^. Indeed, CD8+ T cell infiltration in the kidneys correlates with disease activity and poor renal outcomes in patients with lupus nephritis ^28^. Interestingly, a correlation between the percentage of HLA-DR+ CD8+ T cells in the peripheral blood with the degree of CD8+ T cell infiltration in renal tissues has been reported in patients with lupus nephritis ^28^.

The expression of HLA-DRB1, a class II MHC molecule, on CD8+ T cells is atypical and suggests a state of chronic activation or altered differentiation. While class II expression has been reported on CD8+ T cells in the setting of viral infections, its functional significance remains incompletely defined. It has been previously suggested that class II expression on CD8+ T cells might be explained by trogocytosis, a process whereby CD8+ T cells acquires HLA-class II molecules via physically removing fragments of the plasma membrane from antigen presenting cells ^29^. However, our current transcriptional data and our previous findings demonstrating significant hypomethylation in the *HLA-DRB1* locus in lupus CD8+ T cells suggests intrinsic upregulation of HLA-DRB1 expression ^15^.

Our findings reveal that the HLA-DRB1+ subset in SLE is largely composed of effector memory (EM), terminally differentiated effector memory re-expressing CD45RA (TEMRA), and proliferative cells, consistent with a state of chronic antigen exposure and immune activation. These cells also demonstrate elevated expression of cytolytic genes such as *GZMB*, *PRF1*, and *GNLY*, as well as the inflammatory cytokine *IFNG*, suggesting retained or heightened cytotoxic capacity.

Paradoxically, despite this apparent cytotoxic potential, CD8+ HLA-DRB1+ T cells in SLE also exhibit features of exhaustion and immunosenescence. We observed increased expression of exhaustion markers (*LAG3*, *TIGIT*), upregulation of senescence-related genes (*CDKN2A*, *KLRG1*), and downregulation of co-stimulatory molecules (*CD27*, *CD28*). This dual phenotype reflects a dysfunctional T cell state commonly seen in chronic viral infections and cancer, and increasingly recognized in autoimmune diseases like SLE ^30, 31^. These findings raise the possibility that CD8+ HLA-DRB1+ T cells may be less effective at pathogen control or tissue surveillance, while still contributing to inflammatory damage in lupus-affected organs.

When we compared CD8+ HLA-DRB1+ T cells from SLE patients to those from healthy controls, we observed further skewing toward dysfunction in the lupus context, with significantly reduced cytotoxicity, higher exhaustion scores, and increased expression of immunoregulatory or stress-related genes, including *ENTPD1*, *TNFAIP3*, *EZH2*, and *XIST*. These findings support the concept that chronic immune activation in SLE may reprogram CD8+ T cells into a state that is simultaneously inflammatory, proliferative, and functionally impaired.

Functionally impaired CD8+ T cells with defective cytotoxicity have been linked to increased susceptibility to viral infections in lupus^13^. Mechanistically, this impairment is driven by increased EZH2 activity ^12^, whose expression in our study was significantly higher in CD8+ HLA-DRB1+ T cells from lupus patients compared to healthy controls.

Type I interferons, especially IFN-α, are central mediators of SLE pathogenesis ^32^. In our study, IFN-α stimulation enhanced cytotoxic gene expression and *IFNG* production in CD8+ HLA-DRB1+ T cells from SLE patients but also exacerbated exhaustion and senescence-associated gene programs. Notably, *CDKN1A* and *CDKN2A* were significantly upregulated, and co-stimulatory molecule expression was further diminished. These findings suggest that IFN-α drives a terminal activation state in CD8+ T cells, potentially contributing to their dysfunctional persistence in SLE. The enrichment of IFN-related pathways (e.g., NOD-like receptor signaling, RIG-I signaling, apoptosis, and necroptosis) further underscores the dominant influence of type I IFNs on this T cell subset in lupus.

Our study also sheds light on the broader CD8+ T cell compartment in SLE. We observed a global shift away from naïve toward EM and proliferative cells. This skewed differentiation landscape may reflect chronic antigenic stimulation and type I IFN exposure. Upon IFN-α treatment, even CD8+ HLA-DRB1- T cell subsets showed increased exhaustion, supporting the notion that IFN-driven immune remodeling is not restricted to the HLA-DRB1+ population. However, unlike in CD8+ HLA-DRB1+ T cells, type I interferon attenuated cytotoxicity in CD8+ HLA-DRB1- T cells. This paradoxical effect of type I interferon on cytotoxicity programs in CD8+ T cells warrants further mechanistic evaluation.

Taken together, these data suggest that CD8+ HLA-DRB1+ T cells represent a unique and functionally significant population in lupus. Their paradoxical phenotype, combining cytotoxic activation with exhaustion and senescence, may underlie both pathogenic inflammation and impaired immune regulation in SLE. The chronic IFN-α milieu in lupus appears to drive and sustain this dysfunctional state, highlighting the potential therapeutic avenue by targeting type I IFN signaling or T cell exhaustion in the disease.

Future studies should include longitudinal data to assess the stability or dynamics of the CD8+ HLA-DRB1+ T cell subset over time and with disease activity, and explore the antigen specificity of these cells, their tissue-homing potential, and their exact contribution to organ-specific pathology in lupus.

## Conclusion

Our findings define a distinctive IFN-driven subset of CD8+ T cells in SLE that integrates features of activation, cytotoxicity, exhaustion, and senescence. These results enhance our understanding of CD8+ T cell dysfunction in lupus and may inform the development of targeted therapies to restore immune balance in this complex autoimmune disease.

## Data Availability

All data produced in the present work are contained in the manuscript

## Acknowledgements

This work was supported by the National Institute of Allergy and Infectious Diseases (NIAID) of the National Institutes of Health (NIH) grant number R01 AI097134. This research was supported in part by the University of Pittsburgh Center for Research Computing and Data, RRID:SCR_022735, through the resources provided. Specifically, this work used the HTC cluster, which is supported by NIH award number S10OD028483.

## Author contributions

Huizhong Long: Writing – original draft, Performed experiments, Formal analysis, Data curation. Elio Carmona: Writing – review & editing, Data curation. Mikhail G Dozmorov: Formal analysis, Methodology. Amr H Sawalha: Writing – original draft, Supervision, Resources, Methodology, Funding acquisition, Conceptualization.

