## Supplementary Figures for "Type I interferon drives dysfunction of a distinct CD8+ HLA-DRB1+ T cell subset in systemic lupus erythematosus"

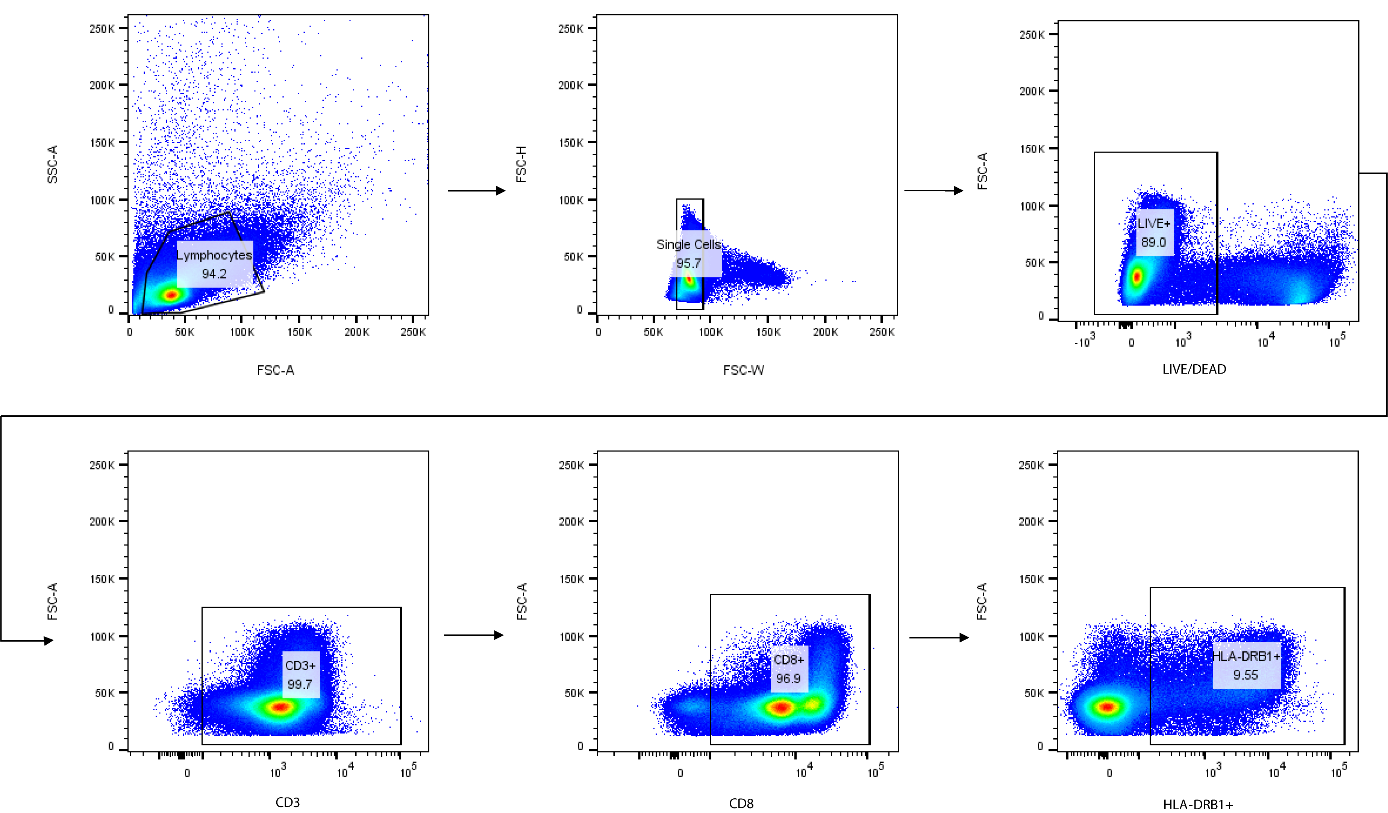

**Supplementary Figure 1: Flow-cytometry gating strategy.** Representative sequential gating used to identify viable CD3+ CD8+ HLA-DRB1+ T cells. Lymphocytes were first selected based on FSC-A vs SSC-A (lymphocyte gate), followed by exclusion of doublets by gating single cells on FSC-H vs FSC-W. Cells were then defined as LIVE/DEAD⁻ events with a viability dye. From the viable singlet lymphocyte population, CD3+ T cells were gated, followed by selection of CD8+ T cells, and finally HLA-DRB1+ cells were quantified within the CD3+ CD8+ compartment (percentages shown on plots indicate the fraction of events within each parent gate).

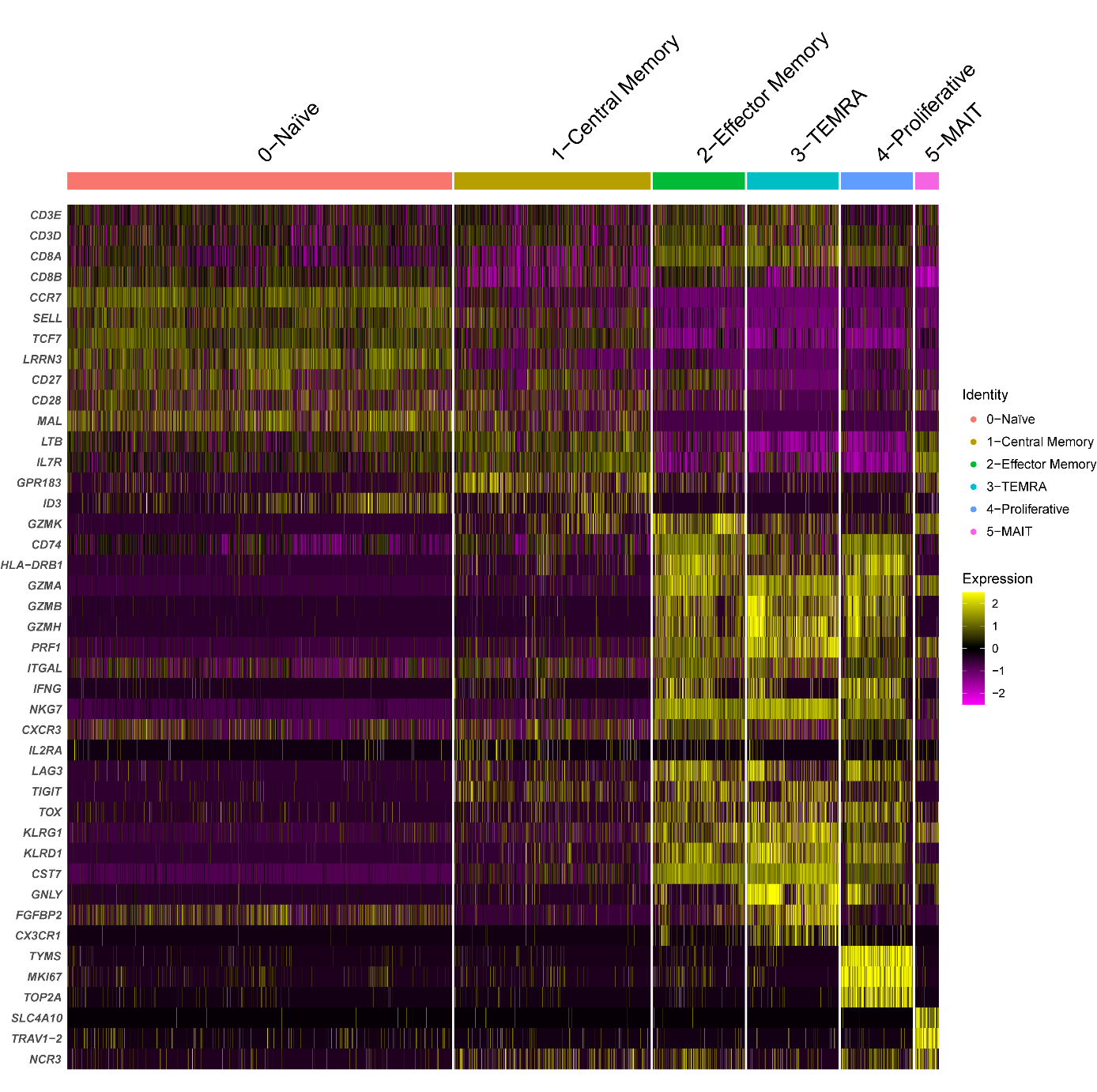

**Supplementary Figure 2:** Heatmap depicting the expression of cell markers in each CD8+ T cell subtype identified using single-cell RNA-seq.

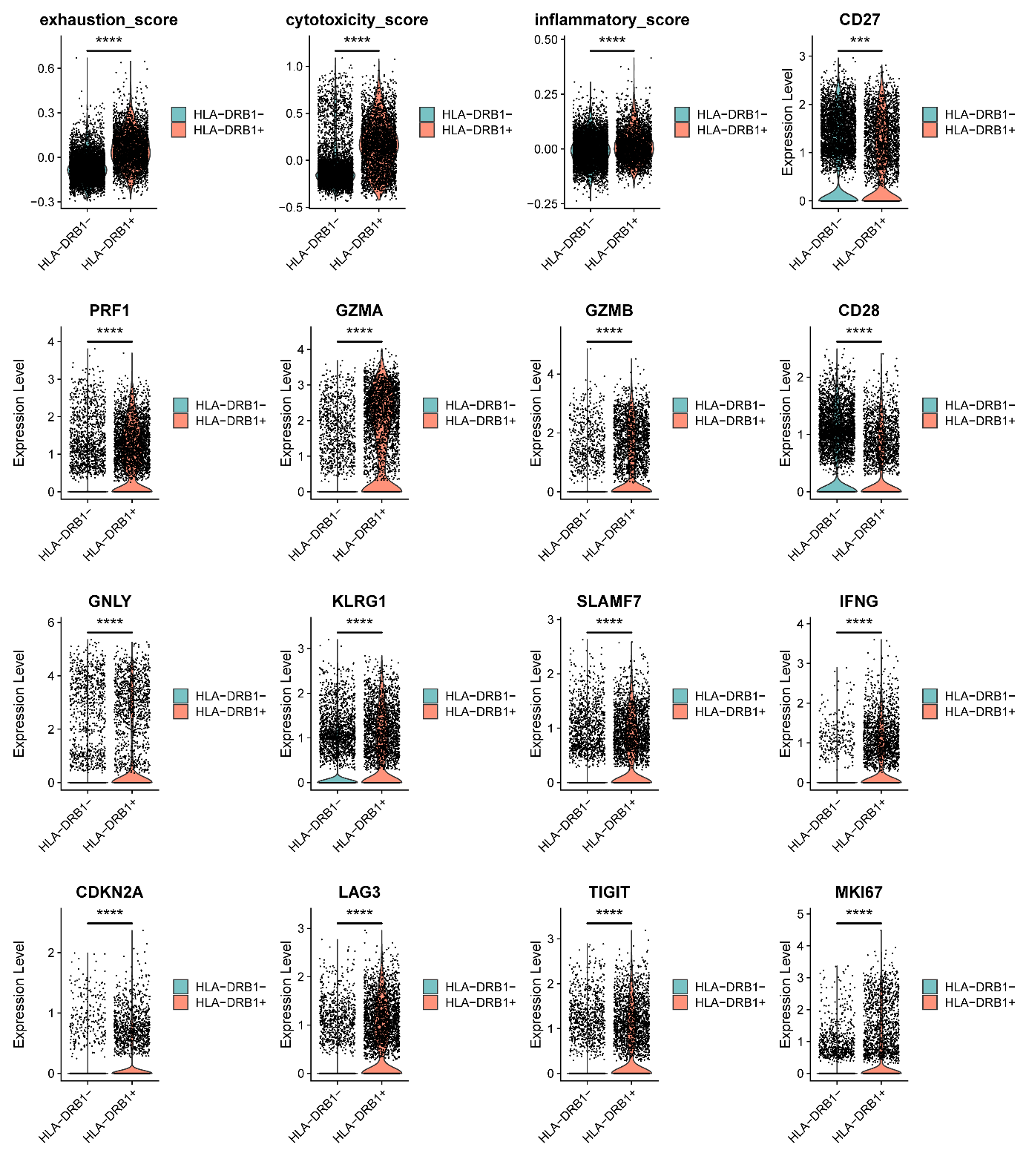

**Supplementary Figure 3: CD8+ HLA-DRB1+ T cells exhibit high cytotoxicity alongside features of exhaustion and senescence.** Violin plots comparing the exhaustion, cytotoxicity, and inflammatory scores (****P < 0.0001, 2-tailed t test), and expression of cytotoxicity, exhaustion, and senescence markers in SLE CD8+HLA-DRB1+ T cells vs CD8+HLA-DRB- T cells (***P < 0.001, ****P < 0.0001; Wilcoxon rank-sum test with FDR correction).

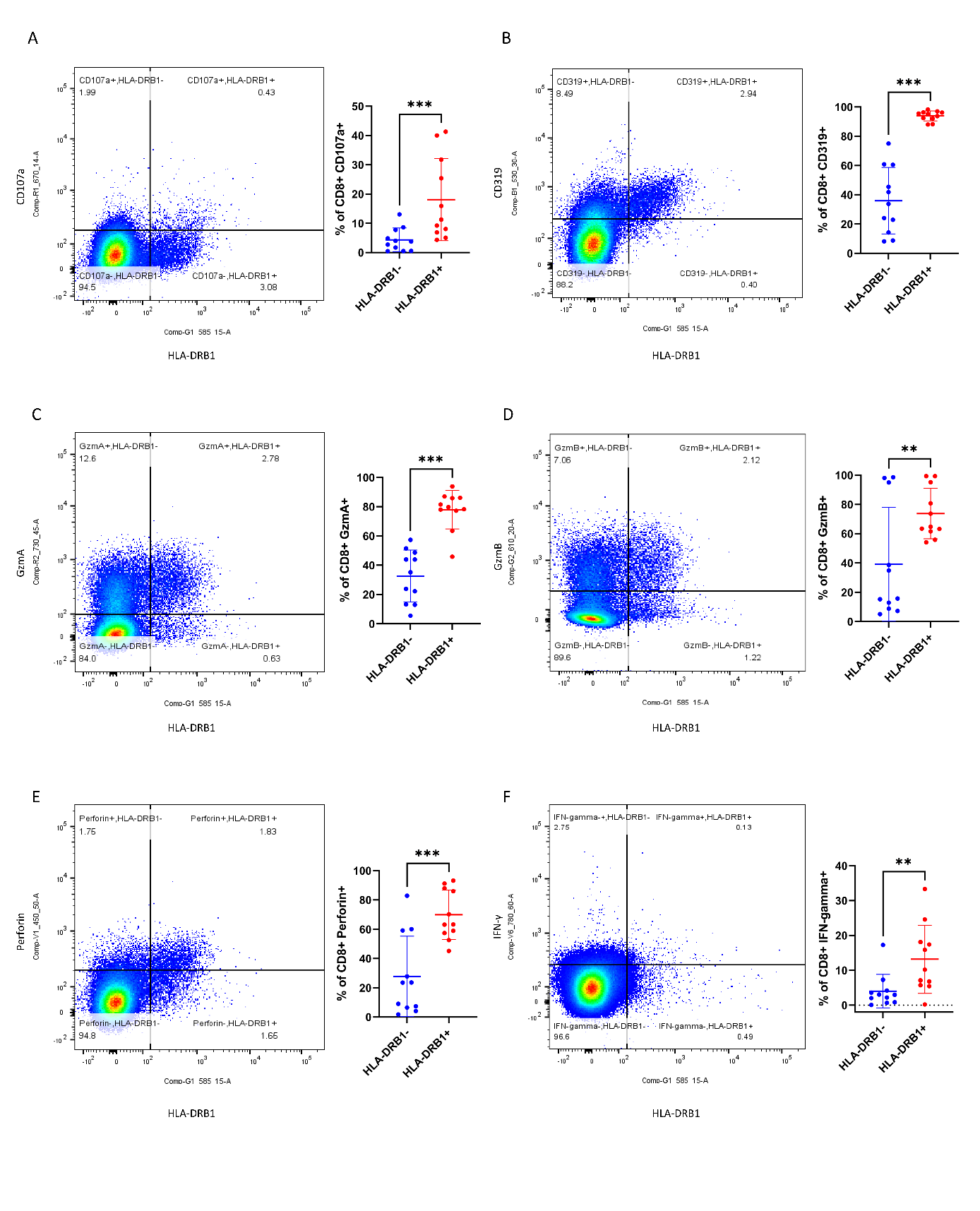

**Supplementary Figure 4:** CD8+ HLA-DRB1+ T cells in patients with SLE demonstrated increased cytotoxicity and inflammatory signatures compared to CD8+ HLA-DRB1- T cells at the protein level, confirming results from single-cell RNA-seq. Representative flow cytometry plots and the proportions of CD107a+ **(A)**, CD319 **(B)**, GzmA **(C)**, GzmB **(D)**, Perforin **(E)**, and IFN-γ **(F)** are shown. **P < 0.01, ***P < 0.001, 2-tailed Mann-Whitney Test.

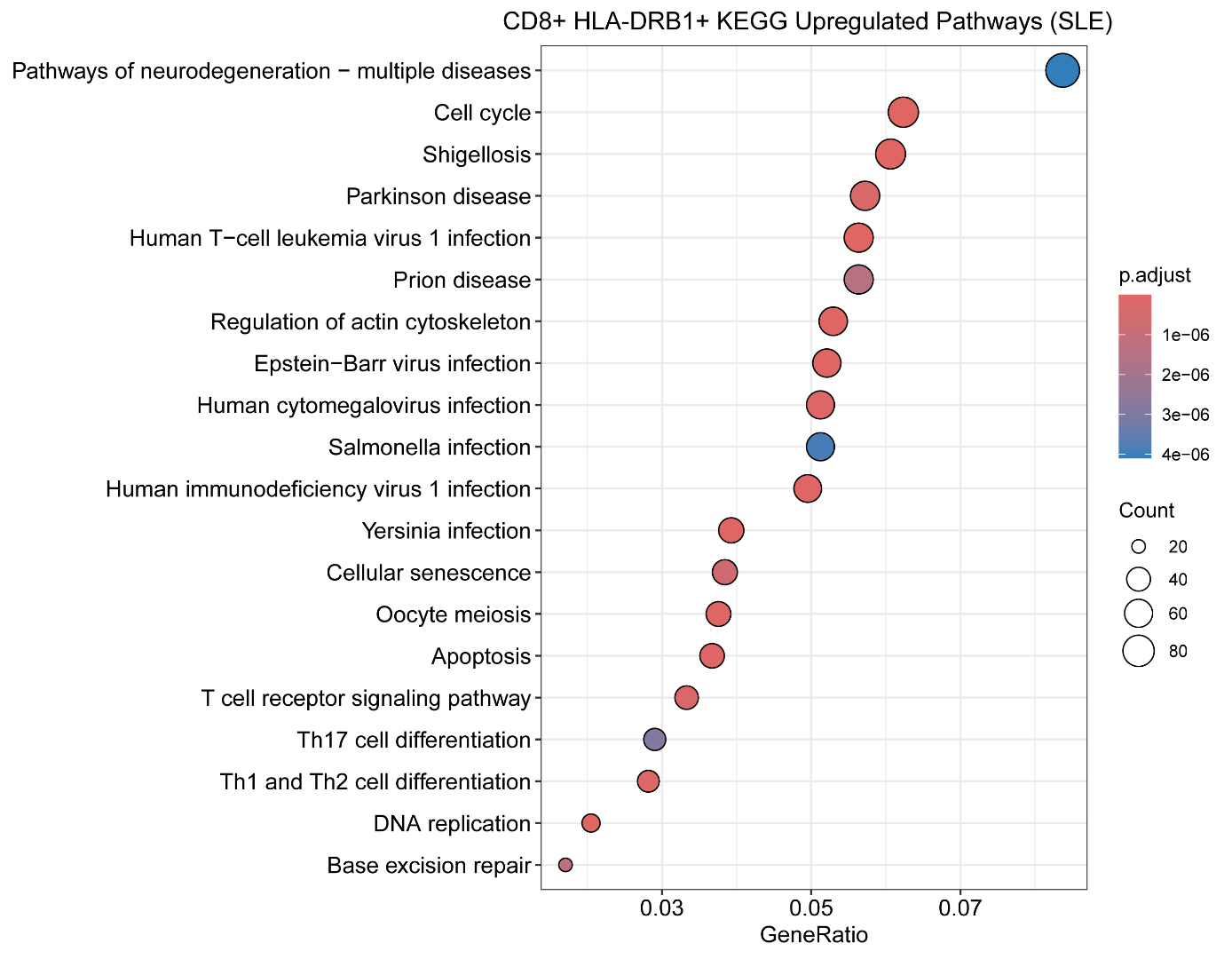

**Supplementary Figure 5:** Representative KEGG terms and pathways enriched in genes upregulated in SLE CD8+ HLA-DRB1+ T cells compared with CD8+ HLA-DRB1- T cells.

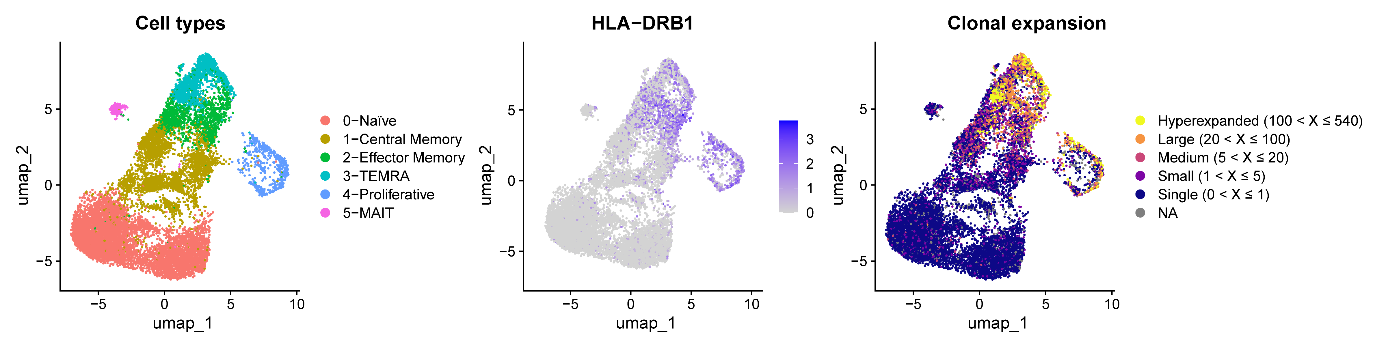

**Supplementary Figure 6: Single-cell RNA-seq profiling of healthy control CD8+T cells after CD3/CD28 stimulation.** **(A)** UMAP plots depicting the cell clusters identified. **(B)** Feature plot depicting the expression of HLA-DRB1. **(C)** UMAP showing the expansion of clonotypes in CD8+ T cells. TCR clonotypes were classified according to their extent of clonal expansion.

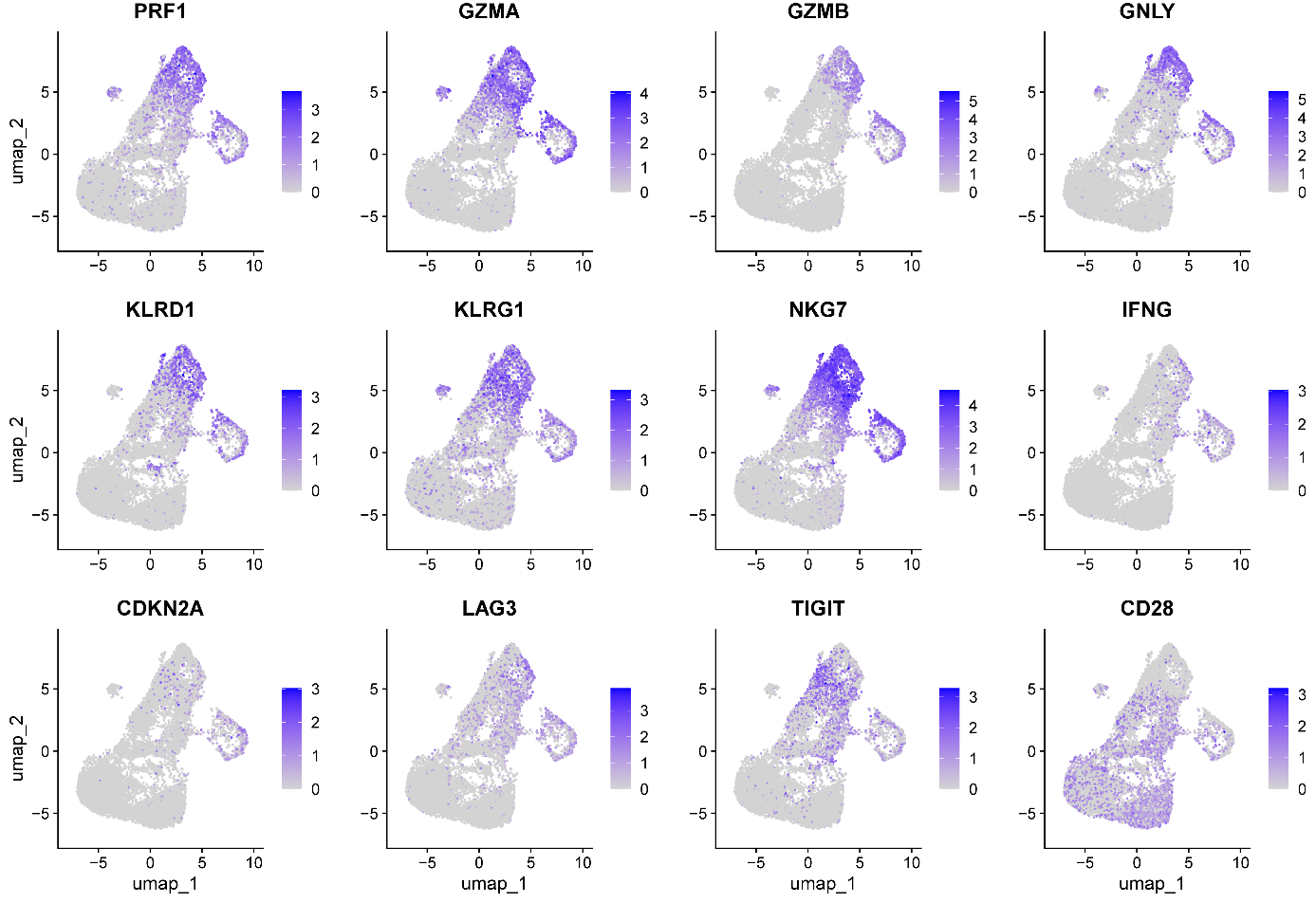

**Supplementary Figure 7: CD8+ HLA-DRB1+ T cells in exhibit high cytotoxicity alongside features of exhaustion and senescence:** Feature plot depicting the expression of cytotoxicity, exhaustion, and senescence markers in healthy control CD8+T cells based on UMAP.

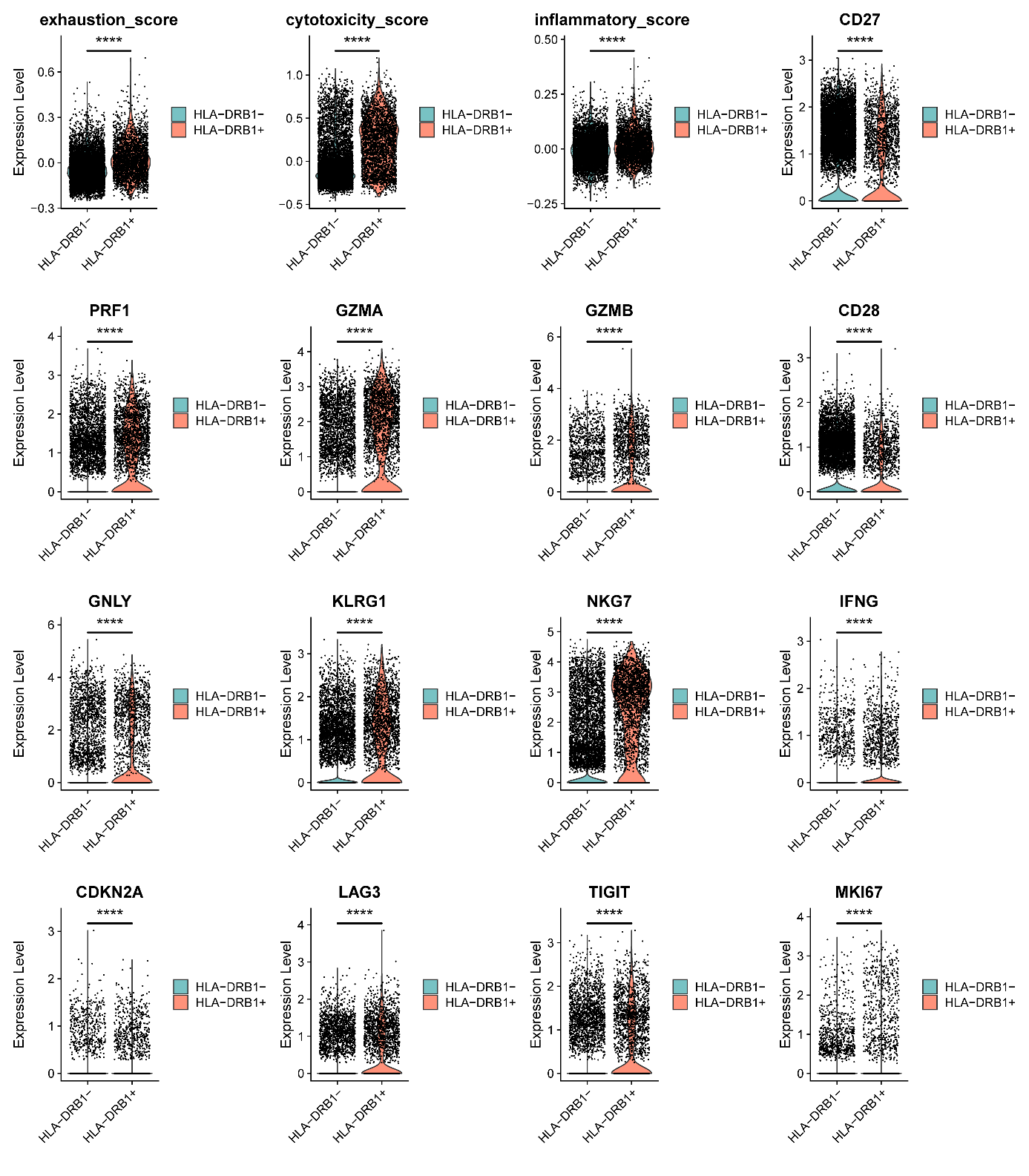

**Supplementary Figure 8: CD8+ HLA-DRB1+ T cells exhibit high cytotoxicity alongside features of exhaustion and senescence.** Violin plots comparing the exhaustion, cytotoxicity, and inflammatory scores (****P < 0.0001, 2-tailed t test), and expression of cytotoxicity, exhaustion and senescence markers in healthy control CD8+ HLA-DRB1+ T cells vs. CD8+ HLA-DRB- T cells (****P < 0.0001, Wilcoxon rank-sum test with FDR correction).

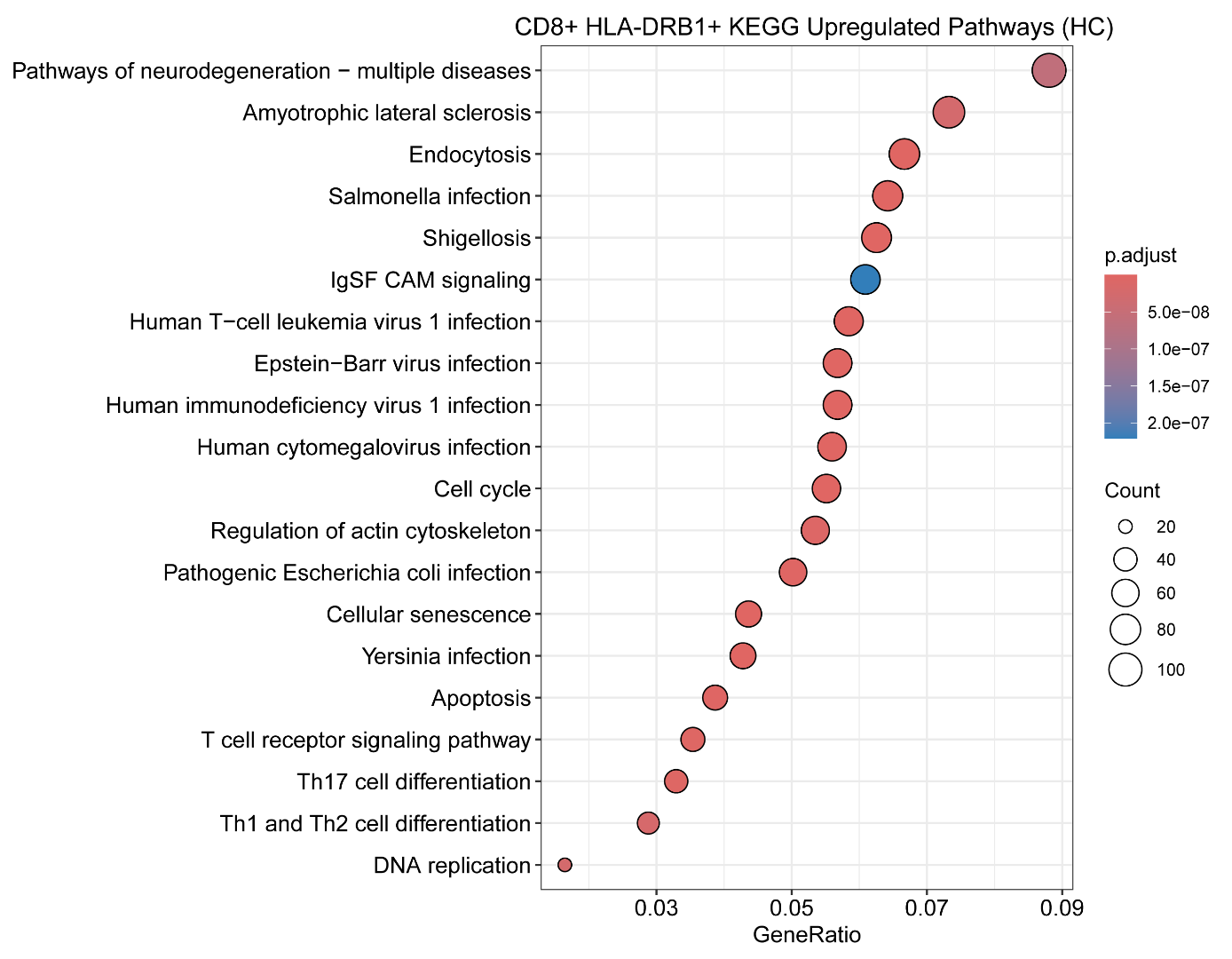

**Supplementary Figure 9:** Representative KEGG terms and pathways enriched in genes upregulated in healthy control CD8+ HLA-DRB1+ T cells compared with CD8+ HLA-DRB1- T cells.

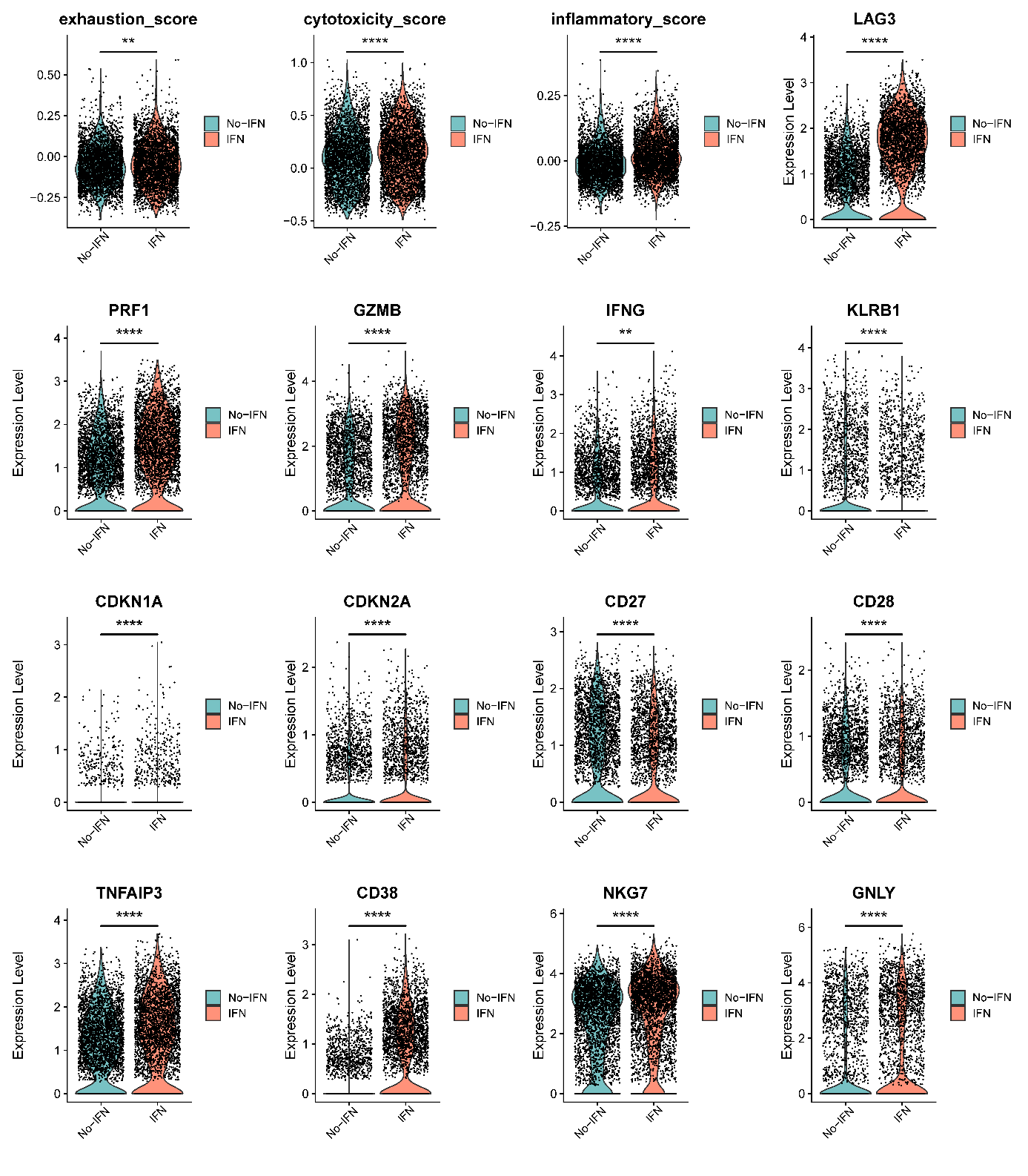

**Supplementary Figure 10: IFN-α treatment enhances the cytotoxicity of SLE CD8+HLA-DRB1+ T cells but also promotes their transition toward exhausted and senescent phenotypes.** Violin plots comparing the exhaustion, cytotoxicity, and inflammatory scores (**P < 0.01, ****P < 0.0001, 2-tailed t test), as well as expression levels of key cytotoxicity, exhaustion, and senescence markers in SLE CD8+HLA-DRB1+ T cells with and without IFN-α treatment (**P < 0.01, ****P < 0.0001, Wilcoxon rank-sum test with FDR correction).

A

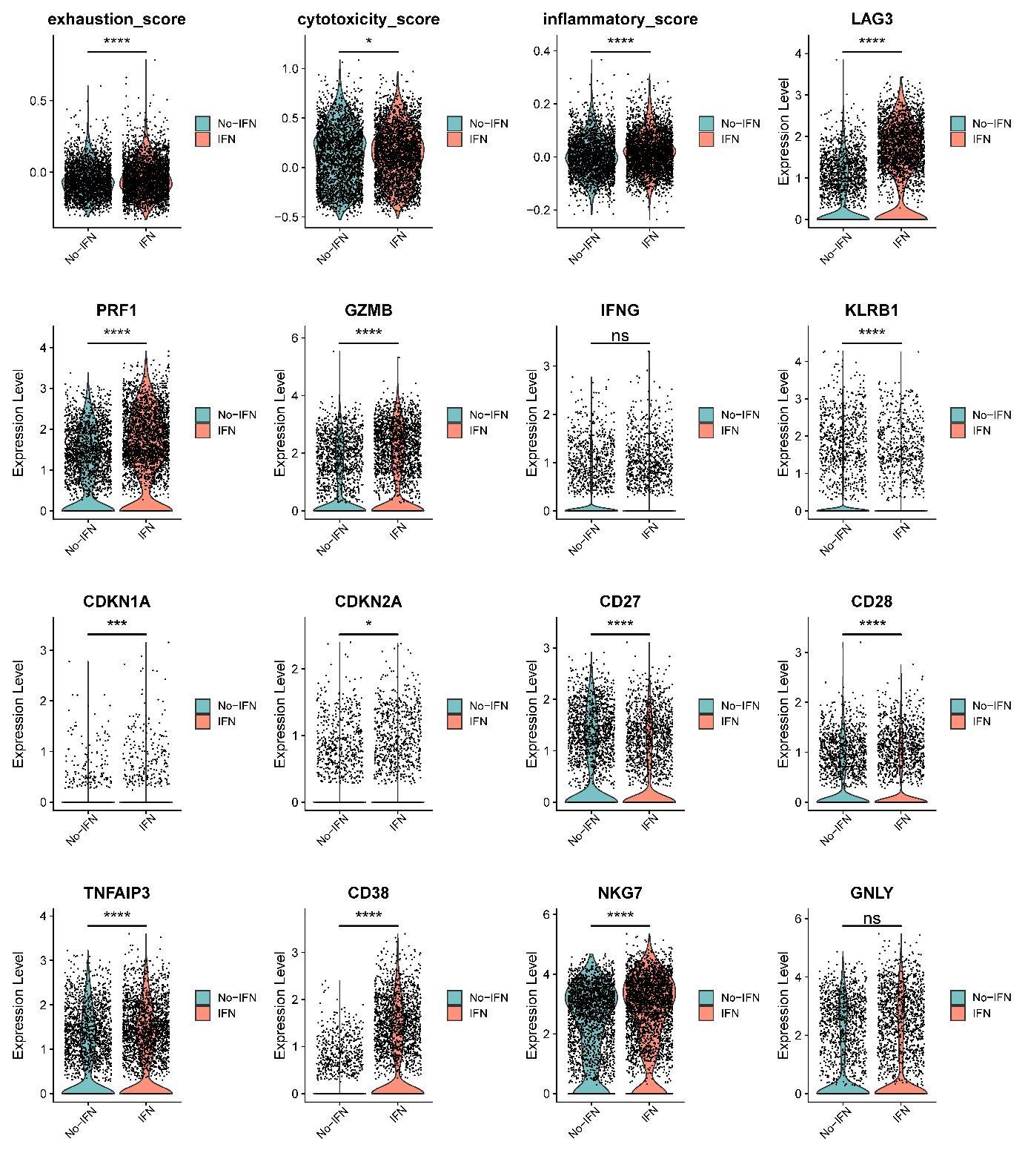

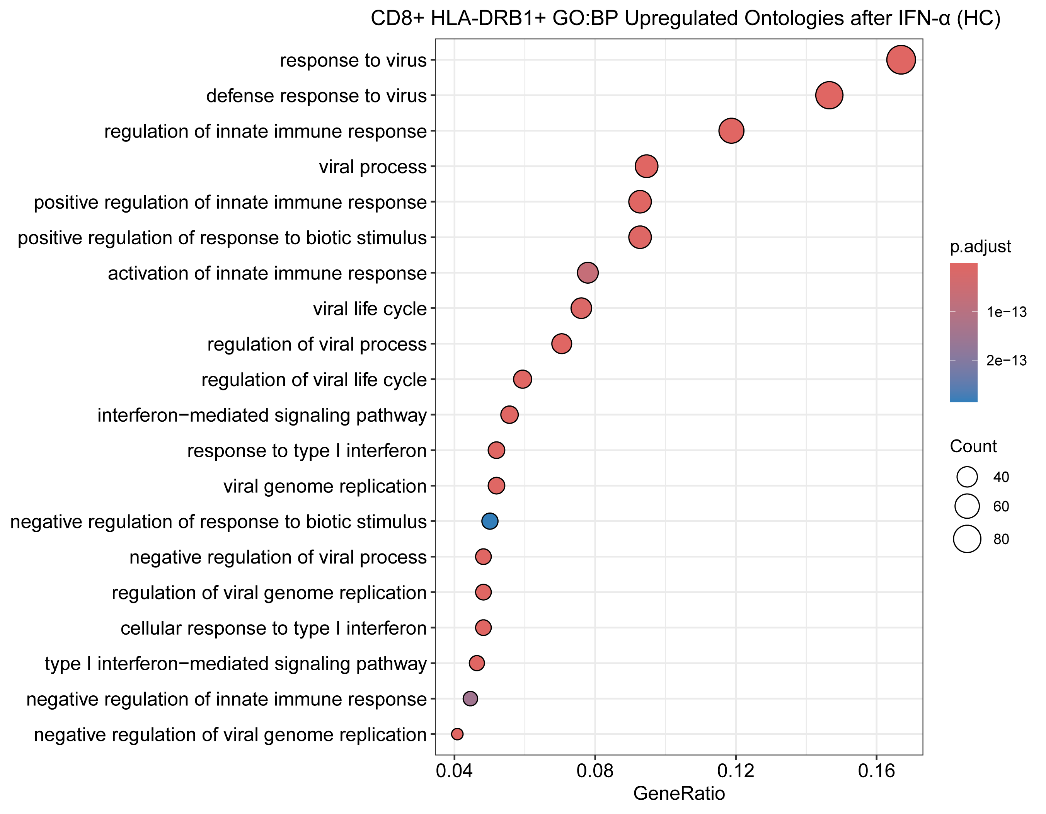
B

C

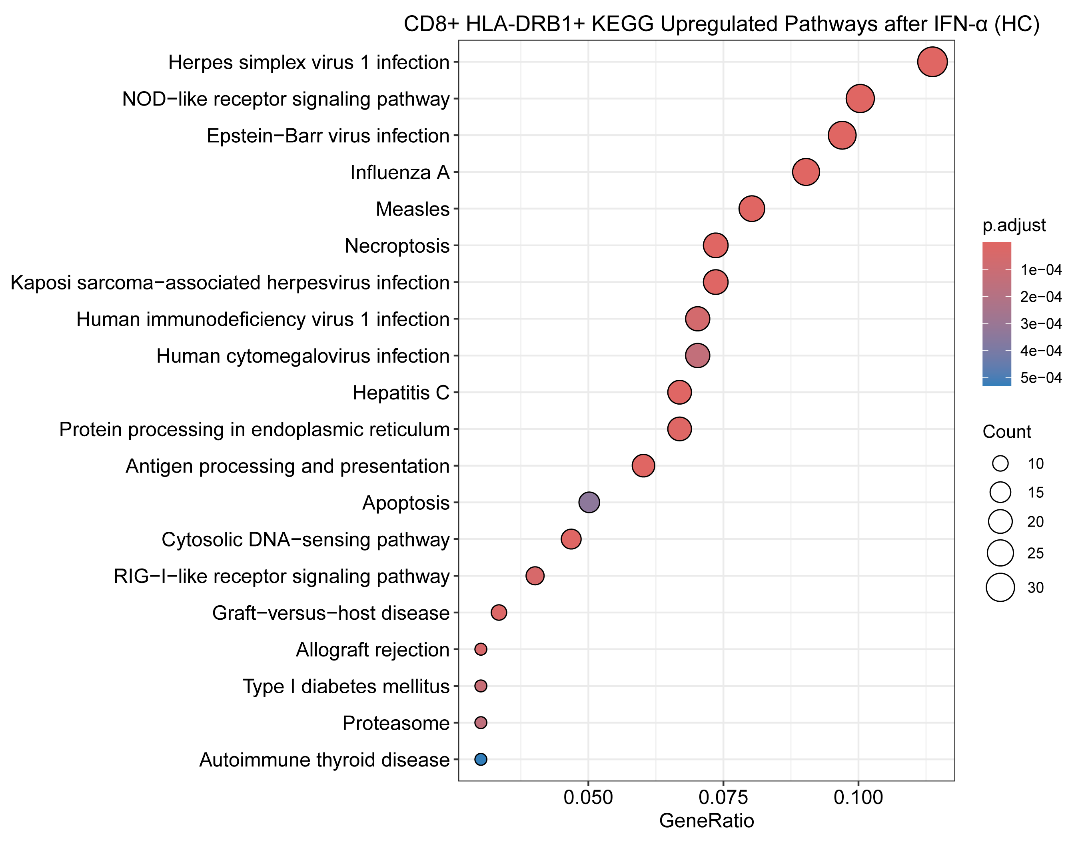

**Supplementary Figure 11: IFN-α treatment enhances the cytotoxicity of healthy control CD8+ HLA-DRB1+ T cells but also promotes their transition toward exhausted and senescent phenotypes. (A)** Violin plots comparing the exhaustion, cytotoxicity, and inflammatory scores (*P < 0.05, ***P < 0.001, ****P < 0.0001, 2-tailed t test), and expression of cytotoxicity, exhaustion and senescence markers in healthy control CD8+ HLA-DRB1+ T cells with and without IFN-α treatment. (ns = not significant, *P < 0.05, **P < 0.01, ****P < 0.0001, Wilcoxon rank-sum test with FDR correction). **(B)** Representative Gene Ontology (Biological Process) terms enriched in genes upregulated after IFN-α treatment in healthy control CD8+ HLA-DRB1+ T cells. **(C)** Representative KEGG terms and pathways enriched in genes upregulated after IFN-α treatment in healthy control CD8+ HLA-DRB1+ T cells.

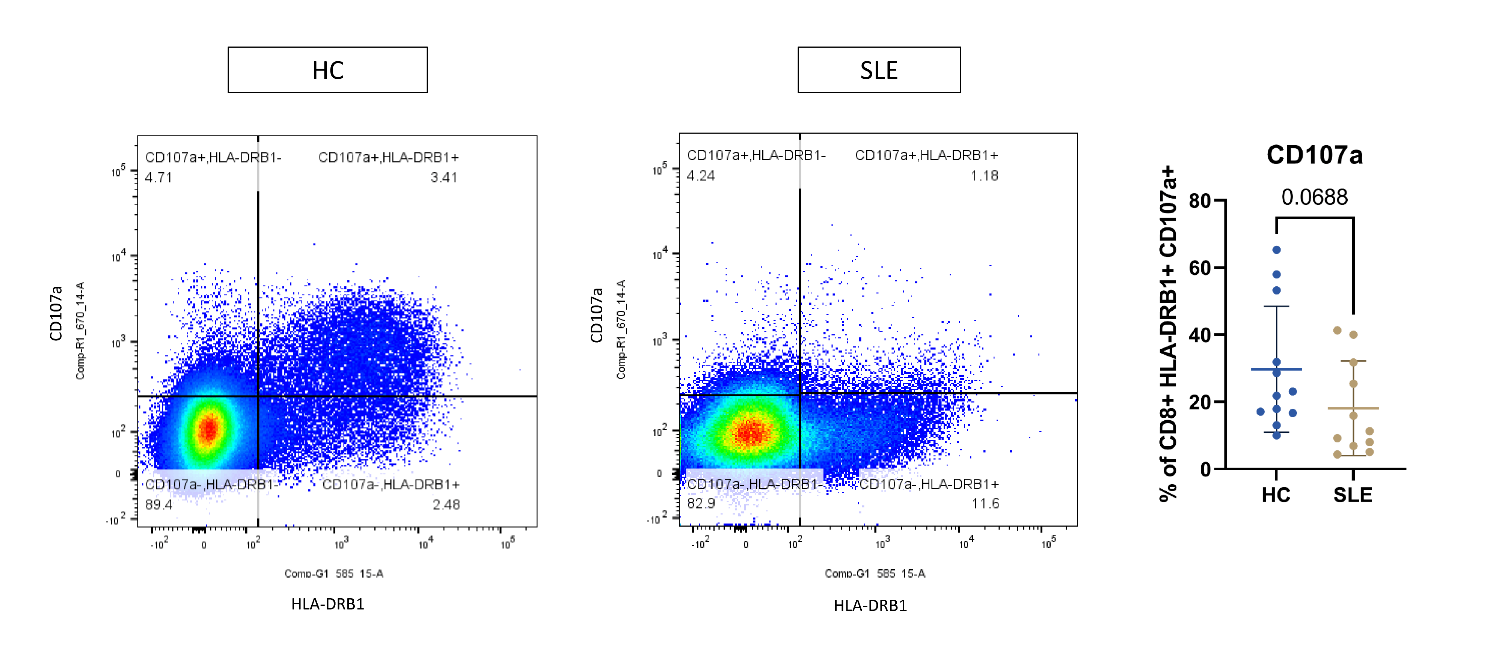

**Supplementary Figure 12**: Reduced CD107a expression in CD8+ HLA-DRB1+ T cells of patients with SLE compared to HLA-DRB1+ T cells in healthy control at the protein level indicative of impaired degranulation. Representative flow cytometry plot and the proportions of CD107a+ are shown. 2-tailed Mann-Whitney Test.

A

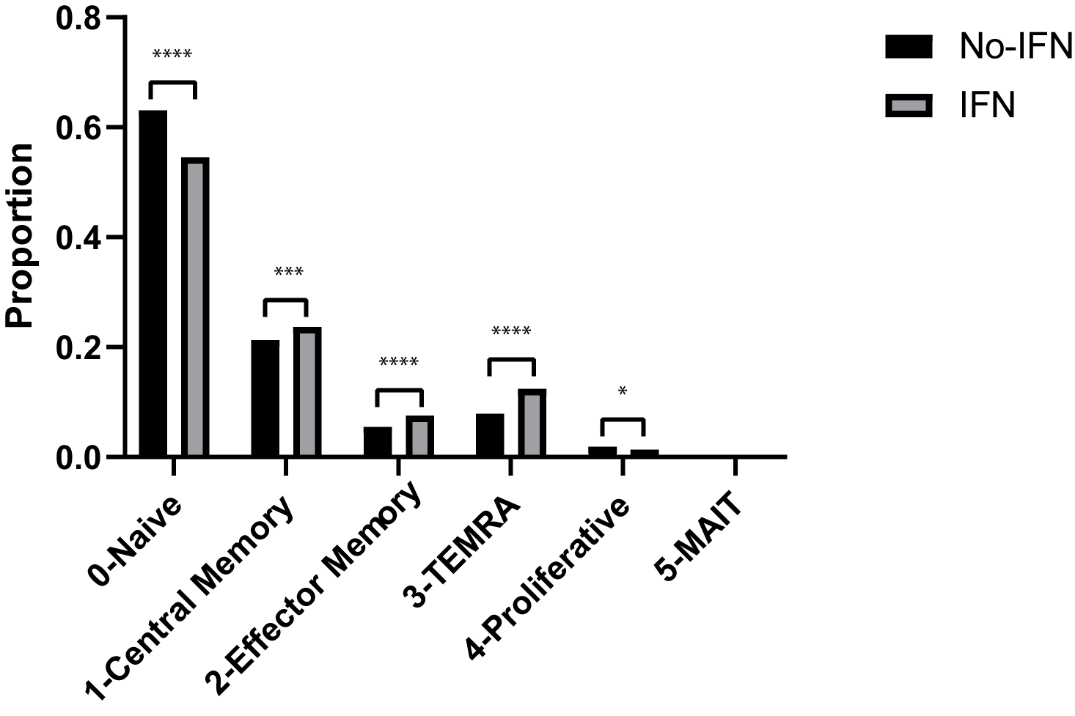

B

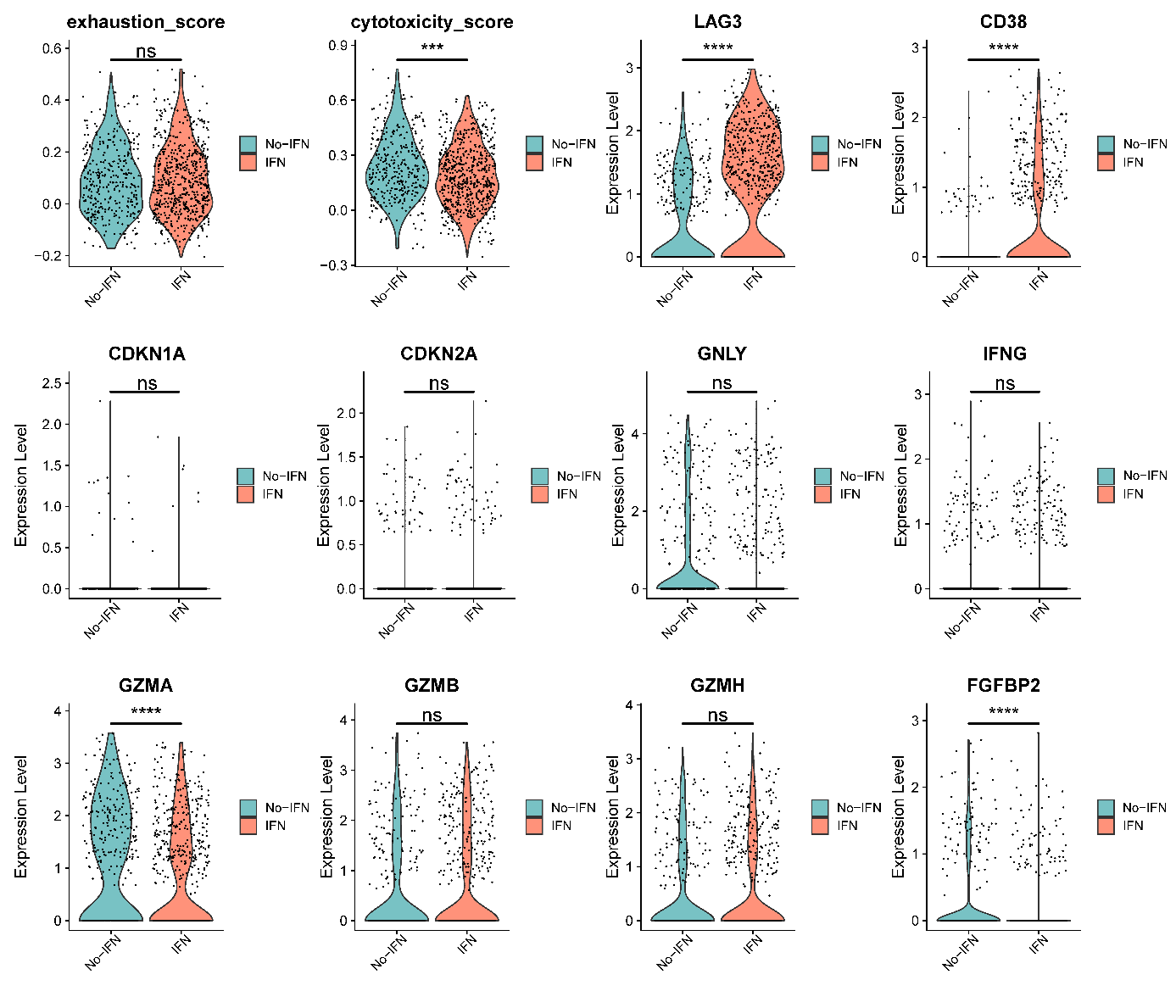

C

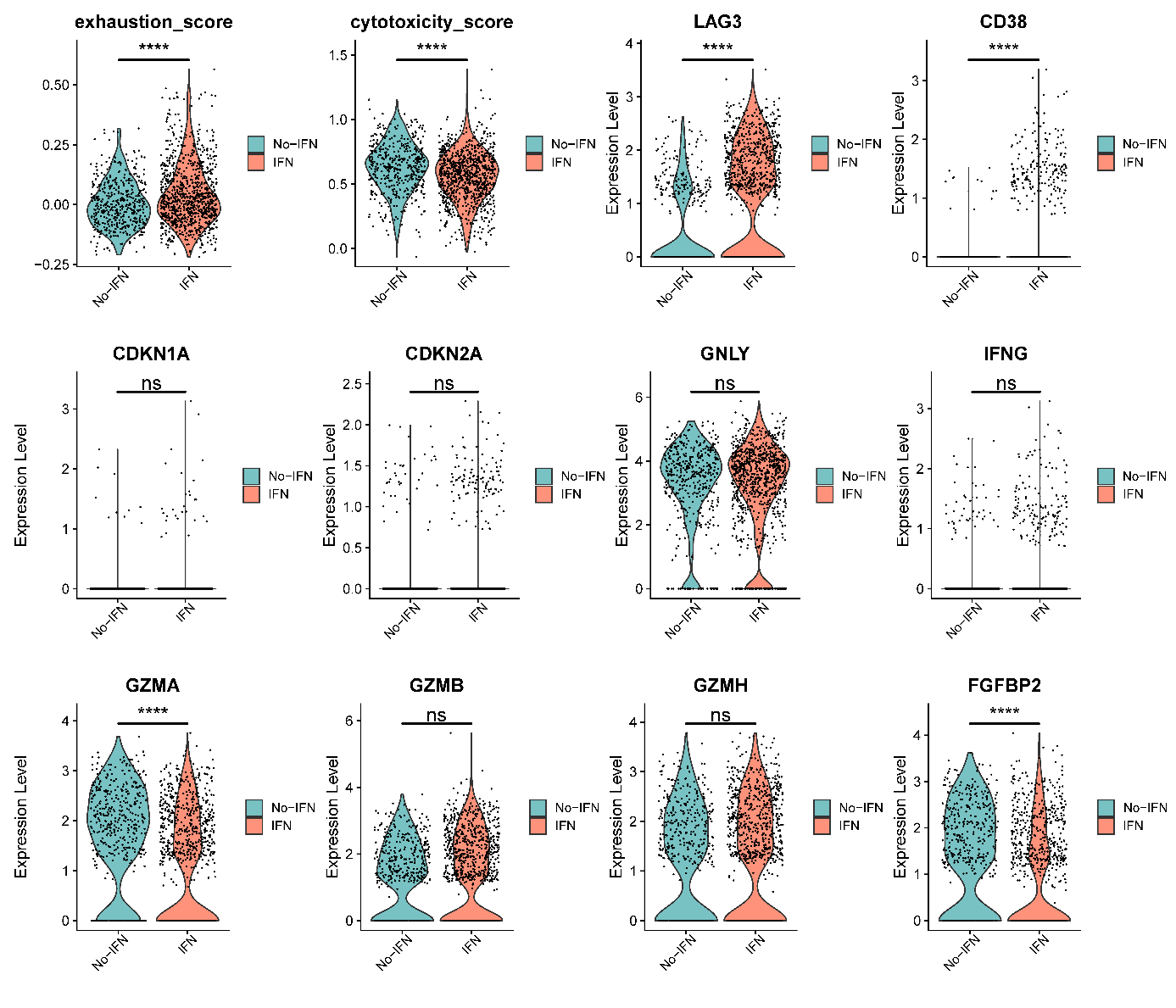

**Supplementary Figure 13: Effect of IFN-α on SLE CD8+ HLA-DRB1- T cells. (A)** Differences in the proportions of SLE CD8+ HLA-DRB1- T cell subtypes with and without IFN-α treatment. ****P < 0.0001, Chi-squared test. **(B)** Violin plots comparing the exhaustion and cytotoxicity scores(ns = not significant, ***P < 0.001, ****P < 0.0001, 2-tailed t test), and expression of cytotoxicity, exhaustion, and senescence markers in SLE CD8+ HLA-DRB1- effector memory (EM) T cells with and without IFN-α treatment (ns = not significant,*P < 0.05, ****P < 0.0001, Wilcoxon rank-sum test with FDR correction). **(C)** Violin plots comparing the exhaustion and cytotoxicity scores (****P < 0.0001, 2-tailed t test), and expression of cytotoxicity, exhaustion, and senescence markers in SLE CD8+ HLA-DRB1- TEMRA cells with and without IFN-α treatment (ns = not significant, ****P < 0.0001, Wilcoxon rank-sum test with FDR correction).

A

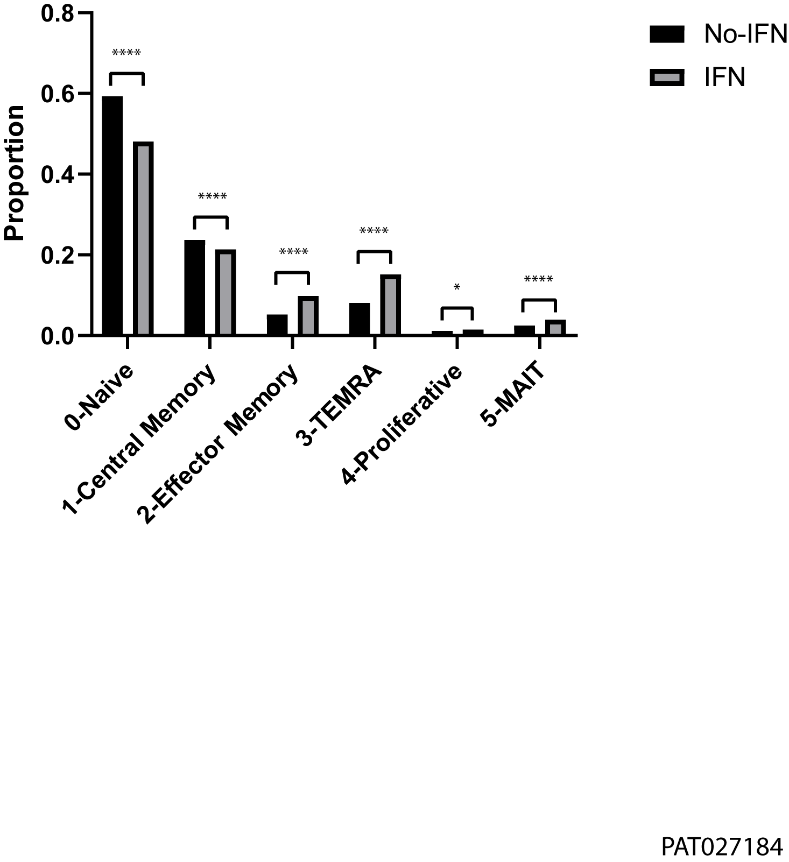

B

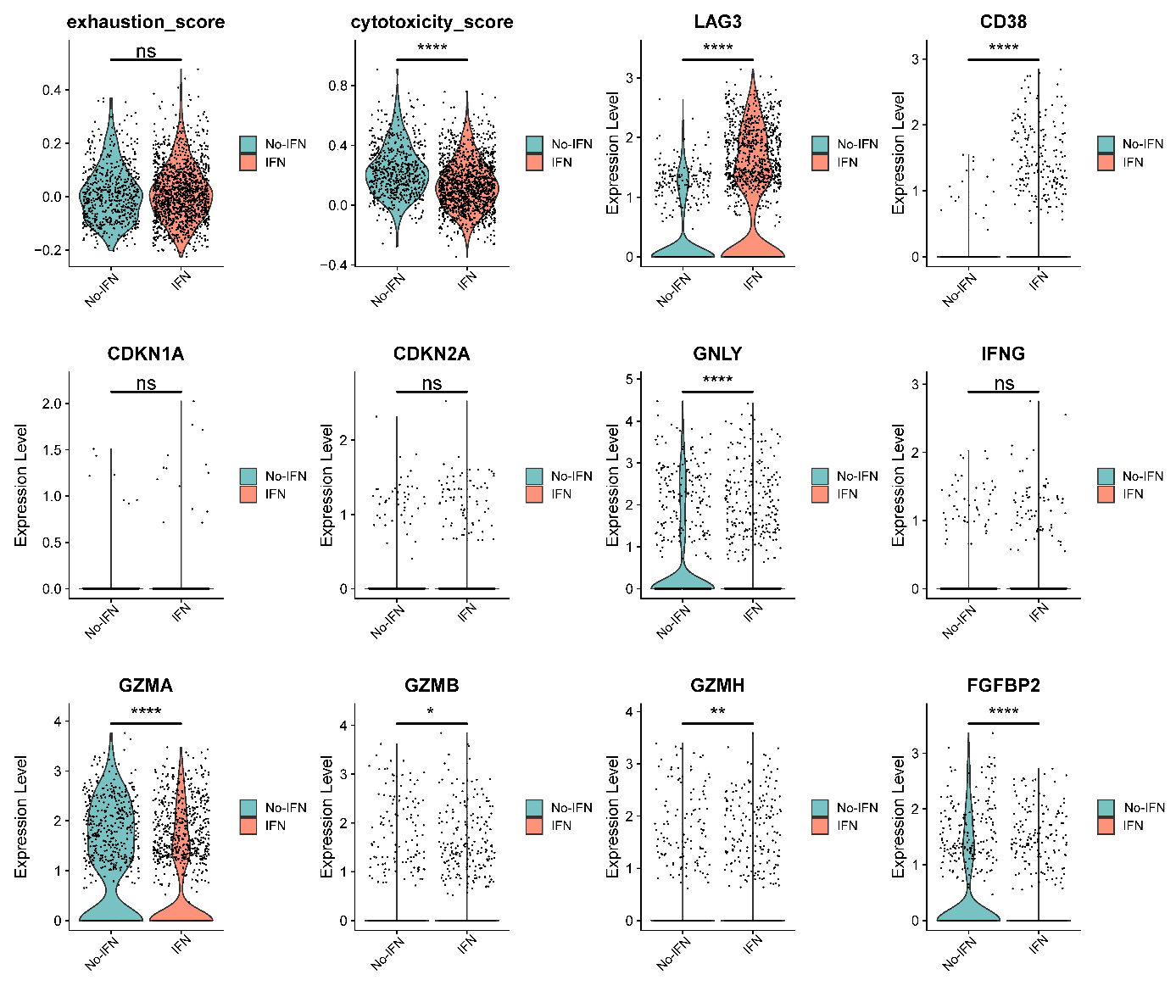

C

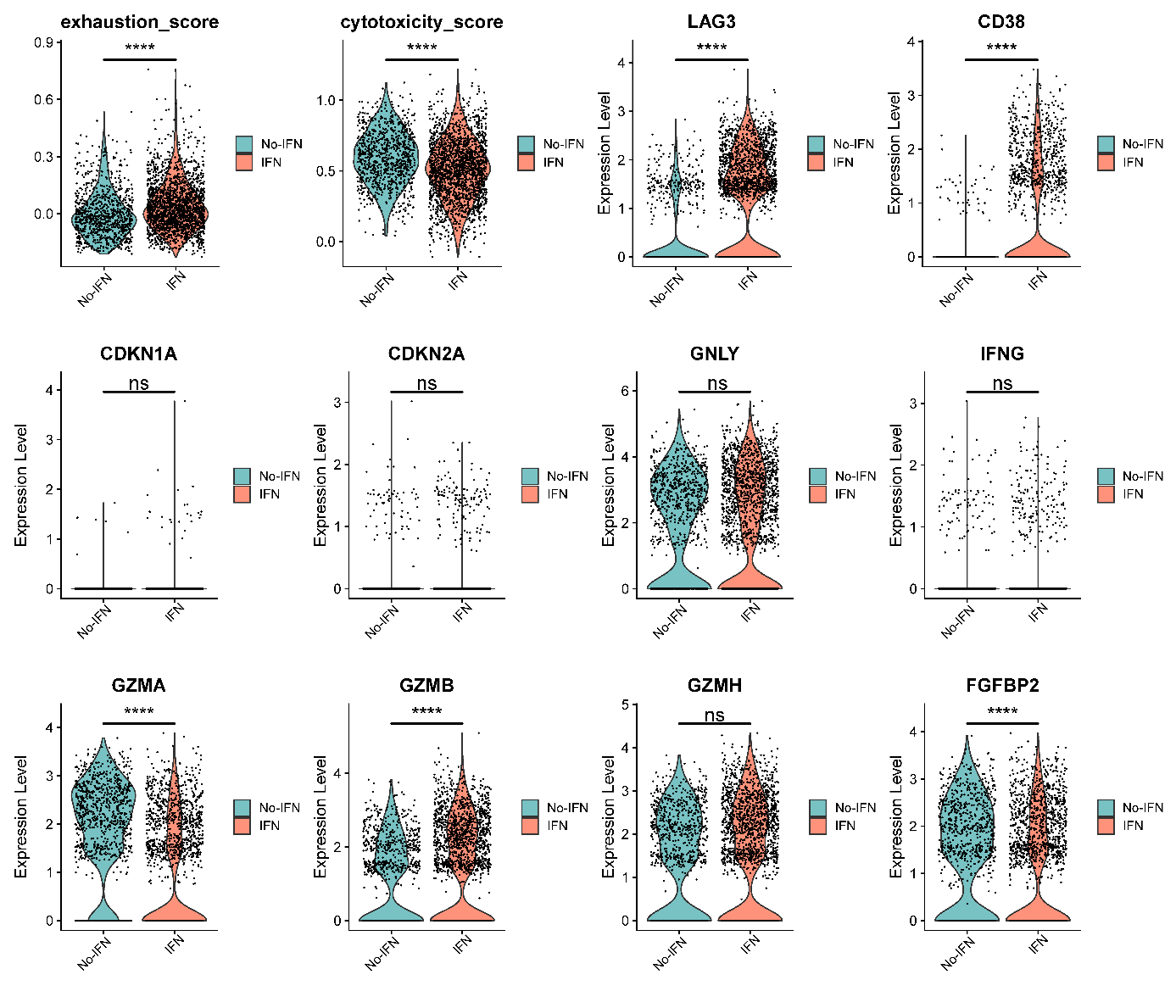

**Supplementary Figure 14: Effect of IFN-α on healthy control CD8+ HLA-DRB1- T cells. (A)** Differences in the proportions of healthy control CD8+ HLA-DRB1- T cell subtypes with and without IFN-α treatment. ****P < 0.0001, Chi-squared test. **(B)** Violin plots comparing the exhaustion and cytotoxicity scores (****P < 0.0001, 2-tailed t test), and expression of cytotoxicity, exhaustion, and senescence markers in healthy control CD8+ HLA-DRB1- effector memory (EM) T cells with and without IFN-α treatment. (ns = not significant, *P < 0.05, **P < 0.01, ****P < 0.0001, Wilcoxon rank-sum test with FDR correction). **(C)** Violin plots comparing the exhaustion and cytotoxicity scores (****P < 0.0001, 2-tailed t test), and expression of cytotoxicity, exhaustion, and senescence markers in healthy control CD8+ HLA-DRB1- TEMRA cells with and without IFN-α treatment (ns = not significant, ****P < 0.0001, Wilcoxon rank-sum test with FDR correction).

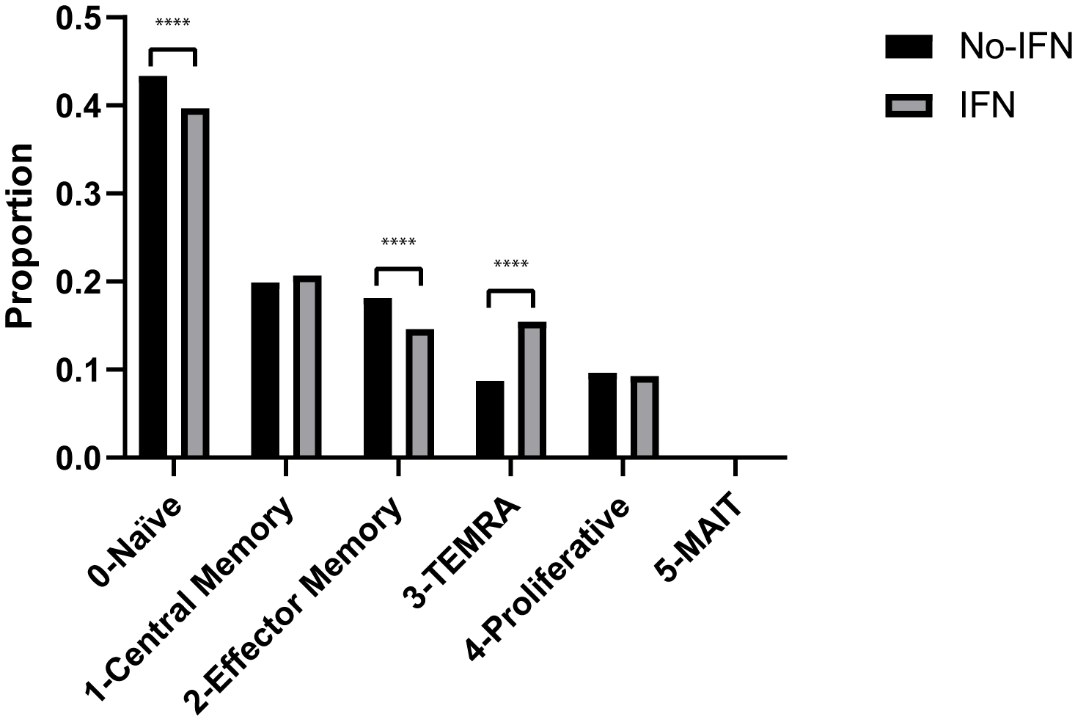

**Supplementary Figure 15:** Differences in the proportions of cell subtypes in SLE CD8+ T cells with and without IFN-α treatment. ****P < 0.0001, Chi-squared test.

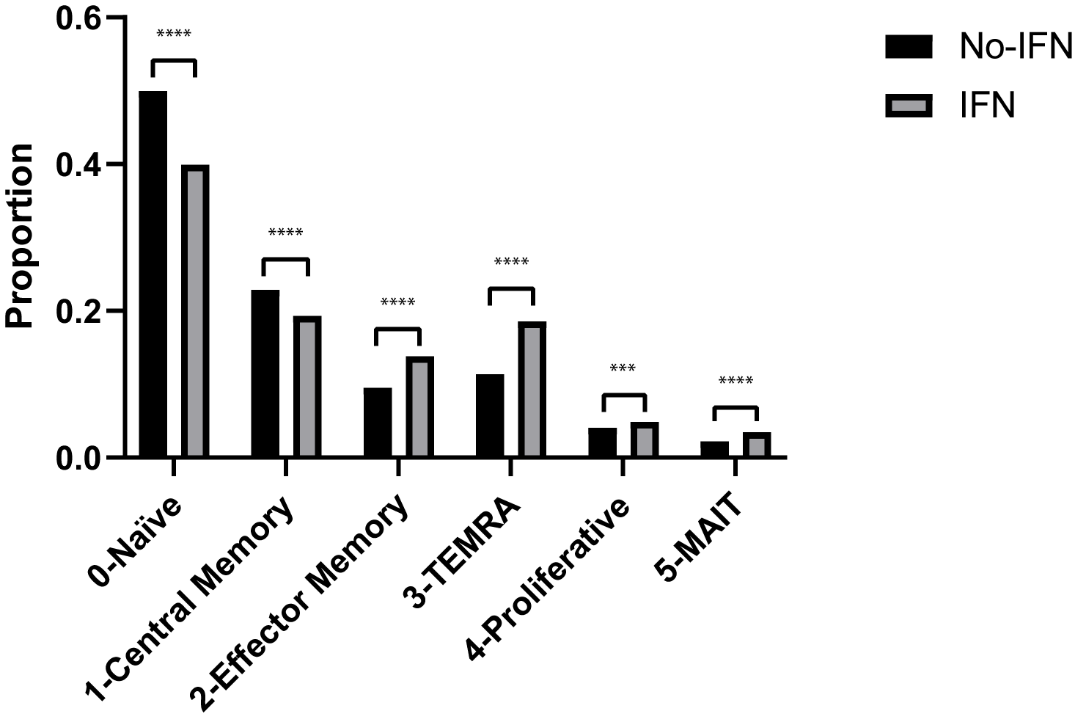

**Supplementary Figure 16:** Differences in the proportions of cell subtypes in healthy control CD8+ T cells with and without IFN-α treatment. ***P < 0.001, ****P < 0.0001, Chi-squared test.

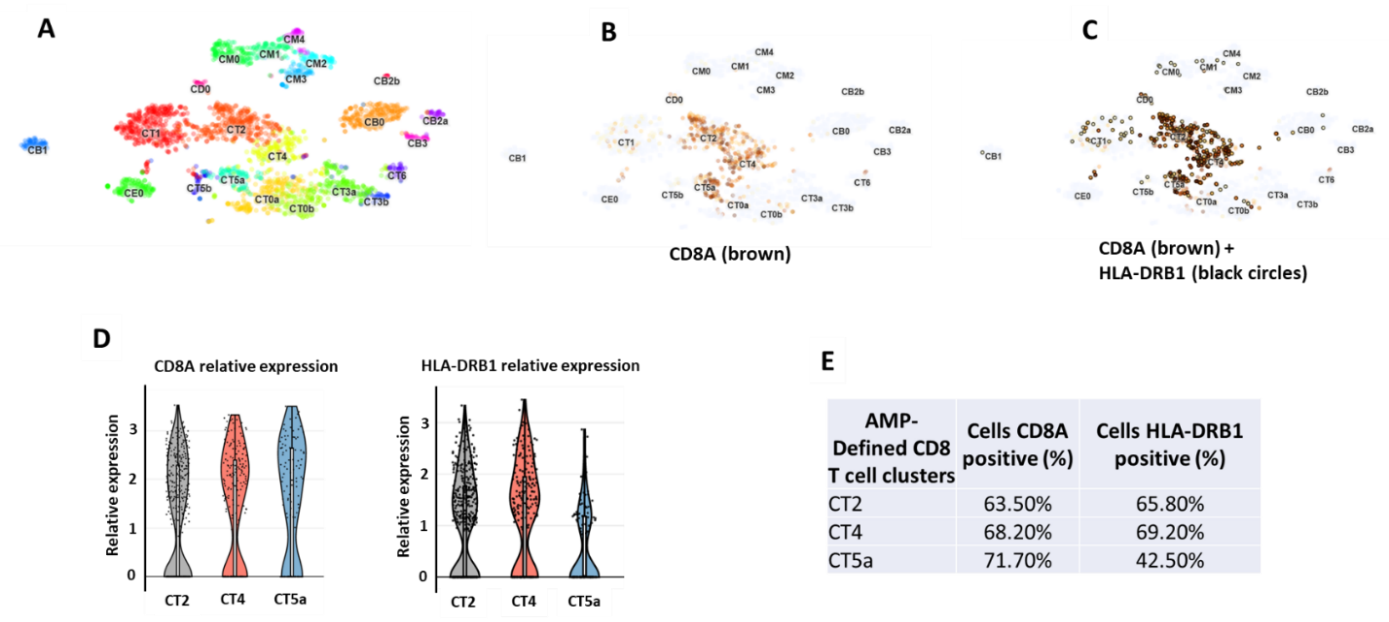

**Supplementary Figure 17: The AMP dataset revealed expression of HLA-DRB1 in the majority of kidney-infiltrating CD8+ T cells. (A)** TSNE plot generated from unsupervised clustering analysis of single cell RNA-seq data from kidney biopsies of lupus patients generated by the AMP project. **(B)** Feature plot showing CD8A expression, revealing 3 clusters of CD8+ T cells as identified by the AMP project (CT2, CT4, and CT5a). **(C)** Feature plot revealing that majority of kidney infiltrating CD8+ T cells in lupus patients also express HLA-DRB1. **(D)** Violin plots showing relative expression of CD8A and HLA-DRB1 in the 3 kidney infiltrating CD8+ T cell clusters in lupus patients. **(E)** Percentage of CD8A and HLA-DRB1 expressing cells in the 3 clusters enriched in CD8+ T cells.
